# Availability and Quality of Anthropometric Data in Swiss Children’s Hospitals: The SwissPedGrowth Project

**DOI:** 10.64898/2026.03.27.26349493

**Authors:** Lorenz M. Leuenberger, Yara Shoman, Franco Romero, Xeni Deligianni, Anna Hartung, Rebeca Mozun, Nicole Goebel, Julia A. Bielicki, Marie-Anne Burkhardt, Philipp Latzin, Christoph Saner, Klara M. Posfay-Barbe, Valérie Schwitzgebel, Eric Giannoni, Michael Hauschild, Martin Stocker, Franziska Righini Grunder, Roger Lauener, Pascal Müller, Luregn J. Schlapbach, Oskar G. Jenni, Ben D. Spycher, Claudia E. Kuehni, Fabiën N. Belle, the SwissPedHealth consortium

## Abstract

**OBJECTIVE:** Anthropometric data are critical in paediatric care, routinely assessed during clinical visits, and available in electronic health records (EHRs). We describe the feasibility of extracting anthropometric data from heterogeneous EHR systems of Swiss children’s hospitals, evaluate their availability and quality, and assess the cohort’s representativeness of the general population.

**METHODS:** In this multicentre study (SwissPedGrowth), we retrospectively collected EHRs from patients <20 years who visited hospitals in Basel, Bern, Geneva, Lausanne, Luzern, St. Gallen, or Zurich between 2017–2023. Sociodemographic, administrative, and clinical information from EHRs were provided in a standardized way by a paediatric national data stream (SwissPedHealth), including the Swiss Neighbourhood Index of Socioeconomic Position (Swiss-SEP). We counted anthropometric recordings per visit to describe availability and used a self-developed and an existing (growthcleanr) algorithm to investigate data quality. To assess representativeness, we compared sociodemographic characteristics between SwissPedGrowth and the general paediatric population in Switzerland, computed standardized differences (effect size: 0.2 small, 0.5 medium, 0.8 large), and weighted the study population to reduce differences.

**RESULTS:** We included 477,531 patients and 2,171,633 hospital visits—54% boys, 71% Swiss, mean Swiss-SEP 65 (SD: 11), and median age at visit 6.3 [IQR: 2.3, 11.8] years. Height recordings were available for 20% of the visits, weights for 43%, and head circumferences for 5%, with better availability for inpatient stays than outpatient or emergency visits. Combining the self-developed and existing algorithm, 4% of heights and 3% of weights were flagged as outliers and 29% of heights and 31% of weights as carried forward from previous visits or same day duplicates. Sociodemographic differences between SwissPedGrowth and the general population were small or small-to-medium and disappeared after weighting.

**CONCLUSION:** SwissPedGrowth demonstrates feasibility of extracting high-quality anthropometric data for paediatric growth research, but challenges regarding completeness and harmonization of EHR data across Swiss hospitals remain.

## 1 INTRODUCTION

Anthropometric measurements, including height, weight, and head circumference are key to paediatric care. Paediatricians use them to monitor growth and development, diagnose overweight, obesity, and short stature, interpret diagnostic tests, and dose medications. Growth patterns deviating from references may indicate underlying diseases, making reliable anthropometric reference data essential for both individual patient care and epidemiological research answering public health questions [1, 2].

Paediatricians routinely collect height and weight measurements of children and adolescents during clinical visits and enter them in electronic health records (EHRs). These data potentially allow for large-scale multi-institutional research networks and longitudinal analyses [3, 4]. The transition from clinical documentation in EHRs to data, which is ready to use for research, is challenging. Barriers may include uncertainties about the availability and quality of recorded measurements [5, 6], technical difficulties of data extraction and harmonization across heterogeneous EHR systems [7, 8], and concerns about representativeness of clinical cohorts relative to the general population [9]. In Switzerland, the SwissPedData and SwissPedHealth initiatives have established a framework to overcome these technical hurdles by enabling harmonized extraction EHR data across Swiss children’s hospitals [10-12]. The potential for paediatric growth research of these large-scale multi-institutional EHR data from SwissPedHealth and their representativeness has not yet been assessed.

In this study, we evaluated the feasibility of extracting anthropometric measurements alongside sociodemographic and clinical information from heterogeneous EHR systems of children’s hospitals, using the SwissPedHealth framework. We investigated data availability and quality and assessed how well the patients represent the demographic characteristics of the paediatric population in Switzerland.

## 2 MATERIALS AND METHODS

### 2.1 Study design

The SwissPedGrowth project is nested in the national data stream SwissPedHealth that combines EHRs of seven children’s hospitals in Switzerland [12]: Basel, Bern, Geneva, Lausanne, Lucerne, St. Gallen, and Zurich. Participating hospitals collected clinical data of in- and outpatients between 2017–2023 who were younger than 20 years old at the visit. SwissPedGrowth was approved by the cantonal ethics commission Bern (KEK Bern 2023-00022) and allowed the re-use data of patients who accepted and signed the general hospital consent between 01.01.2017 and 31.12.2025 and of patients who were informed about general consent, and did not actively refuse, between 01.01.2017 and 28.02.2023. For this analysis, we excluded patients with missing sex and visits for which we were unable to define the type of visit, e.g. missing information about the department.

### 2.2 Data extraction and preparation

Hospitals participating in SwissPedHealth extracted sociodemographic, administrative, anthropometric, and clinical data from electronic source systems and loaded them into local clinical data warehouses (CDWs). They linked the Neighbourhood Index of Socioeconomic Position (Swiss-SEP) to the patient’s home address using a standard operating procedure [13]; the patient’s address was deleted after linkage. The Swiss-SEP is an area-based socioeconomic index, including information on education, occupation, rent, and overcrowding, and ranging from 0 (low socioeconomic status) to 100 (high socioeconomic status) [14]. We used the Swiss-SEP version 3, updated with census data from 2012–2015 [15]. CDWs mapped the data to the schema of the Swiss Personalized Health Network (SPHN) using standard terminologies (e.g. SNOMED-CT, LOINC), converted it into a graphical data format with quality control rules, the resource description framework (RDF), and sent it to BioMedIT—a secure server for processing and analysing medical data [16]. We converted (flattened) the RDF data into tabular format for data cleaning and analysis. To create a dataset of body mass index (BMI), for every weight recording we matched the closest height recording allowing for a maximum of 30 days between the recordings (consistent with previous EHR-based growth studies) and calculated BMI as weight/height^2^ [17]. See supplementary methods for detailed information about data extraction and preparation, and for a description of the datasets (supplementary Table 1).

We defined three types of visits: outpatient visit: healthcare encounter with ambulatory care; emergency visit: healthcare encounters to an emergency department; inpatient stay: healthcare encounters with an overnight stay. We grouped nationality as Swiss, European, and non-European. We categorized diagnoses into three categories based on International Classification of Disease 10^th^ version (ICD-10) codes: Affecting growth permanently (e.g. malignant neoplasms [C00-C97]), affecting growth temporarily (e.g. appendicitis [K35-K37]), no diagnoses affecting growth. The list of ICD-10 codes is based on exclusion criteria of previous growth studies and expert opinion (supplementary Table S2) [18, 19].

To describe challenges during data extraction in detail, paediatricians at the participating hospitals manually reviewed EHRs of 70 outpatient and 70 inpatient visits (20 per hospital), randomly selected from visits where height or weight recordings were not extracted.

### 2.3 Statistical analysis

To describe data availability, we counted the number of height, weight, BMI, and head circumference recordings per visit. We stratified the availability of recordings by type of visit, age at visit, and hospital. We compared characteristics between visits with or without an anthropometric recording. We described data availability before excluding any recordings using the data quality assessment below.

To assess data quality of height and weight, we identified unit errors, decimal errors, swapped recordings (height recorded as weight and vice versa), duplicated recordings (measured on the same day, or carried forward from previous visits), biologically implausible outliers, and invalid recordings (zero or negative value, negative age) using three approaches: 1) running a self-developed algorithm based on z-scores of anthropometric measurements—calculated using the WHO growth references adopted to Switzerland [20]—, 2) running the existing growthcleanr [21] algorithm based on a longitudinal moving average of a patient’s z-scores, and 3) running our self-developed and existing algorithms sequentially combined. For head circumference recordings, we used our self-developed algorithm. For calculated BMI values, we used the results of the combined algorithms for the weight and height recordings and additionally flagged biologically implausible BMI values as z-scores <-5 or >8. Details about the algorithms are presented in the supplementary methods.

To assess how well the patients in SwissPedGrowth represent the general population, we compared their age, sex, nationality, and Swiss-SEP to the entire Swiss population below 20 years of age based on census data from the Swiss Federal Statistical Office (FSO) [22]. We calculated standardized mean differences using Glass’ Δ for normally distributed variables and standardized proportion differences using Cohen’s h for categorical variables [23, 24]. We interpreted the effect sizes according to Cohen [24]: 0.2 small, 0.5 medium, 0.8 large. We weighted the SwissPedGrowth patients to match the distributions of age, sex, nationality, and Swiss-SEP of the general population by calculating weights through iterative proportional fitting (raking) using the *survey* package in R [25]. We only applied these weights to verify representativeness, not for analysing the availability and quality of anthropometric data.

We used GraphDB Workbench version 10.8.0 to flatten RDF data and R Studio Server version 2024.03.999 with R version 4.3.1 for the statistical analysis [26-28]. The analysis code is available on GitHub (https://github.com/LorenzLeuenberger/Availability-Quality-SwissPedGrowth).

## 3 RESULTS

### 3.1 Study population

We received data of 560,995 patients and 2,346,460 hospital visits from the SwissPedHealth national data stream. We excluded 83 patients with missing sex (n=83, <1%) and 15% of patients because of missing type of visit (n=83,341, 15%), since both were required for our availability analyses. We included 477,531 (85%) patients and 2,171,633 (93%) visits for analysis (supplementary Figure S1), including 54% boys and 71% of Swiss nationality (Table 1 and supplementary Table S3). The mean Swiss-SEP was 65 (SD: 11).

**Table 1.**
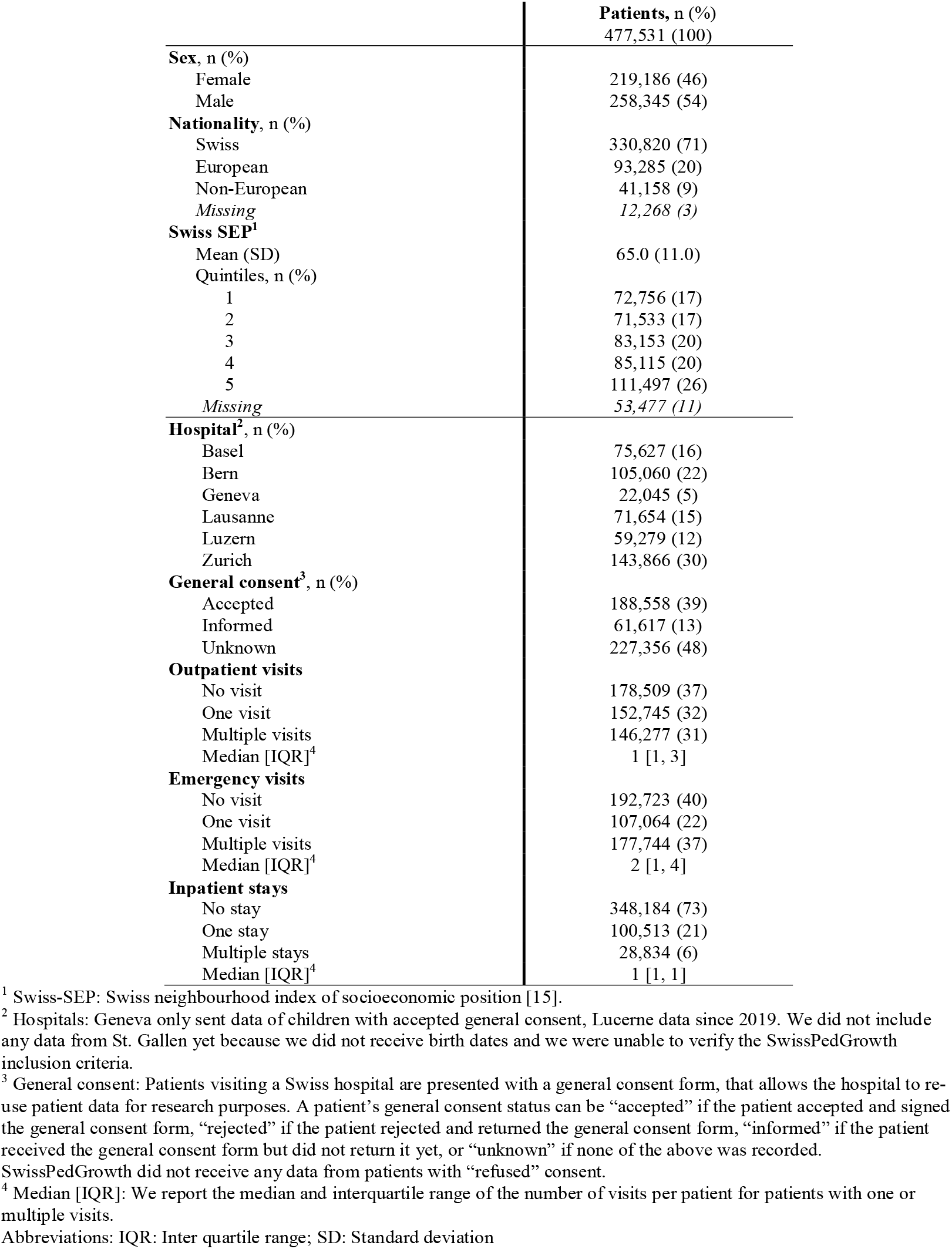
Sociodemographic and administrative information of patients in the SwissPedGrowth project.

|  | Patients, n (%) |
| --- | --- |
|  | 477,531 (100) |
| **Sex, n (%)** |  |
| Female | 219,186 (46) |
| Male | 258,345 (54) |
| **Nationality, n (%)** |  |
| Swiss | 330,820 (71) |
| European | 93,285 (20) |
| Non-European | 41,158 (9) |
| Missing | 12,268 (3) |
| **Swiss SEP<sup>1</sup>** |  |
| Mean (SD) | 65.0 (11.0) |
| Quintiles, n (%) |  |
| 1 | 72,756 (17) |
| 2 | 71,533 (17) |
| 3 | 83,153 (20) |
| 4 | 85,115 (20) |
| 5 | 111,497 (26) |
| Missing | 53,477 (11) |
| **Hospital<sup>2</sup>, n (%)** |  |
| Basel | 75,627 (16) |
| Bern | 105,060 (22) |
| Geneva | 22,045 (5) |
| Lausanne | 71,654 (15) |
| Luzern | 59,279 (12) |
| Zurich | 143,866 (30) |
| **General consent<sup>3</sup>, n (%)** |  |
| Accepted | 188,558 (39) |
| Informed | 61,617 (13) |
| Unknown | 227,356 (48) |
| **Outpatient visits** |  |
| No visit | 178,509 (37) |
| One visit | 152,745 (32) |
| Multiple visits | 146,277 (31) |
| Median [IQR]<sup>4</sup> | 1 [1, 3] |
| **Emergency visits** |  |
| No visit | 192,723 (40) |
| One visit | 107,064 (22) |
| Multiple visits | 177,744 (37) |
| Median [IQR]<sup>4</sup> | 2 [1, 4] |
| **Inpatient stays** |  |
| No stay | 348,184 (73) |
| One stay | 100,513 (21) |
| Multiple stays | 28,834 (6) |
| Median [IQR]<sup>4</sup> | 1 [1, 1] |

<sup>1</sup> Swiss-SEP: Swiss neighbourhood index of socioeconomic position [15].
<sup>2</sup> Hospitals: Geneva only sent data of children with accepted general consent, Lucerne data since 2019. We did not include any data from St. Gallen yet because we did not receive birth dates and we were unable to verify the SwissPedGrowth inclusion criteria.
<sup>3</sup> General consent: Patients visiting a Swiss hospital are presented with a general consent form, that allows the hospital to re-use patient data for research purposes. A patient's general consent status can be "accepted" if the patient accepted and signed the general consent form, "rejected" if the patient rejected and returned the general consent form, "informed" if the patient received the general consent form but did not return it yet, or "unknown" if none of the above was recorded. SwissPedGrowth did not receive any data from patients with "refused" consent.
<sup>4</sup> Median [IQR]: We report the median and interquartile range of the number of visits per patient for patients with one or multiple visits.
Abbreviations: IQR: Inter quartile range; SD: Standard deviation

Patients were a median 6.3 [2.3, 11.8] years old at visit; 59% were outpatient visits, 33% emergency visits, and 9% inpatient stays (Table 2). The median age was higher for outpatient visits (7.9 [3.3, 13.0] years), compared to emergency visits (4.6 [1.8, 9.2]) and inpatient stays (3.6 [0.4, 10.4]). Diagnostic information was available for 81% of inpatient stays, with 26% of them having diagnoses potentially affecting growth permanently, 30% affecting growth temporarily, and 43% having no diagnoses that could affect growth. Diagnostic codes were available for 81% of inpatient stays, but only available for 7% of outpatient and 11% of emergency visits.

**Table 2.** Patient, administrative, and clinical information of hospital visits in the SwissPedGrowth project.

|  | **Total visits, n (%)**<br>2,171,633 (100) | **Outpatient visits, n (%)**<br>1,271,392 (59) | **Emergency visits, n (%)**<br>707,654 (33) | **Inpatient stays, n (%)**<br>192,587 (9) |
| --- | --- | --- | --- | --- |
| **Sex, n (%)** |  |  |  |  |
| Female | 1,203,138 (55) | 706,387 (56) | 389,232 (55) | 107,519 (56) |
| Male | 968,495 (45) | 565,005 (44) | 318,422 (45) | 85,068 (44) |
| **Age at hospital visit (years)** |  |  |  |  |
| Median [IQR] | 6.3 [2.3, 11.8] | 7.9 [3.3, 13.0] | 4.6 [1.8, 9.2] | 3.6 [0.4, 10.4] |
| **Age group, n (%)** |  |  |  |  |
| Infancy: <1 | 302,695 (14) | 138,649 (11) | 101,597 (14) | 62,449 (32) |
| Preschool age: 1–5 | 747,071 (34) | 372,228 (29) | 320,308 (45) | 54,535 (28) |
| School age: 6–11 | 597,336 (28) | 377,707 (30) | 182,722 (26) | 36,907 (19) |
| Early adolescence: 12–15 | 524,531 (24) | 382,808 (30) | 103,027 (15) | 38,696 (20) |
| Late adolescence: 16–19 |  |  |  |  |
| **Nationality, n (%)** |  |  |  |  |
| Swiss | 1,464,135 (69) | 870,919 (70) | 461,562 (67) | 131,654 (70) |
| European | 432,883 (20) | 243,183 (20) | 152,448 (22) | 37,252 (20) |
| Non-European | 222,242 (10) | 129,030 (10) | 74,451 (11) | 18,761 (10) |
| Missing | 52,373 (2) | 28,260 (2) | 19,193 (3) | 4,920 (3) |
| **Swiss SEP<sup>1</sup>** |  |  |  |  |
| Mean (SD) | 64.8 (11.0) | 64.6 (11.0) | 65.3 (11.0) | 64.1 (10.9) |
| **Quintiles, n (%)** |  |  |  |  |
| 1 | 359,536 (18) | 215,222 (18) | 112,394 (17) | 31,920 (19) |
| 2 | 343,490 (17) | 200,211 (17) | 113,808 (17) | 29,471 (18) |
| 3 | 391,695 (20) | 226,499 (19) | 131,887 (20) | 33,309 (20) |
| 4 | 399,832 (20) | 231,758 (20) | 134,769 (20) | 33,305 (20) |
| 5 | 511,747 (26) | 290,406 (25) | 182,438 (27) | 38,903 (23) |
| Missing | 165,333 (8) | 107,296 (8) | 32,358 (5) | 25,679 (13) |
| **Hospital<sup>2</sup>, n (%)** |  |  |  |  |
| Basel | 500,816 (23) | 309,791 (24) | 156,686 (22) | 34,339 (18) |
| Bern | 463,715 (21) | 267,788 (21) | 153,834 (22) | 42,093 (22) |
| Geneva | 254,488 (12) | 162,129 (13) | 77,069 (11) | 15,290 (8) |
| Lausanne | 162,868 (7) | 118,846 (9) | 8,066 (1) | 35,956 (19) |
| Luzern | 209,542 (10) | 108,297 (9) | 83,016 (12) | 18,229 (9) |
| Zurich | 580,204 (27) | 304,541 (24) | 228,983 (32) | 46,680 (24) |
| **General consent<sup>3</sup>, n (%)** |  |  |  |  |
| Accepted | 1,266,437 (58) | 802,465 (63) | 359,057 (51) | 104,915 (54) |
| Informed | 232,293 (11) | 124,792 (10) | 84,792 (12) | 22,709 (12) |
| Unknown | 672,903 (31) | 344,135 (27) | 263,805 (37) | 64,963 (34) |
| **Diagnoses at hospital discharge<sup>4</sup>** |  |  |  |  |
| Affecting growth permanently | 79,297 (24) | 32,287 (34) | 5,607 (7) | 41,403 (26) |
| Affecting growth temporarily | 66,787 (20) | 5,413 (6) | 14,256 (18) | 47,118 (30) |
| No diagnoses affecting growth | 182,310 (56) | 55,938 (60) | 58,487 (75) | 67,885 (43) |
| Missing | 1,843,239 (85) | 1,177,754 (93) | 629,304 (89) | 36,181 (19) |

<sup>1</sup> Swiss-SEP: Swiss neighbourhood index of socioeconomic position [15].
<sup>2</sup> Hospitals: Geneva only sent data of children with accepted general consent, Lucerne data since 2019. We did not include any data from St. Gallen yet because of missing information on birth dates and we were unable to verify the SwissPedGrowth inclusion criteria.
<sup>3</sup> General consent: Patients visiting a Swiss hospital are presented with a general consent form, that allows the hospital to re-use patient data for research purposes. A patient's general consent status can be "accepted" if the patient accepted and signed the general consent form, "rejected" if the patient rejected and returned the general consent form, "informed" if the patient received the general consent form but did not return it yet, or "unknown" if none of the above was recorded. SwissPedGrowth did not receive any data from patients with "refused" consent.
<sup>4</sup> Diagnoses: We categorized diagnoses based on ICD-10 codes or ICD-10 codes associated to ORPHA codes into three categories: Affecting growth permanently, affecting growth temporarily, no diagnoses affecting growth. The list of ICD-10 coded diagnoses and their classification can be found in supplementary Table 1. Many visits did not have diagnostic information because only inpatient visits need to be ICD-10 coded for billing purposes in Switzerland.
Abbreviations: ICD-10: International classification of diseases 10<sup>th</sup> version, IQR: Inter quartile range; ORPHA: Orphanet nomenclature of rare diseases, SD: Standard deviation.

### 3.2 Availability of anthropometric data

We included 675,665 height, 1,747,562 weight, and 192,631 head circumference recordings and calculated 1,169,075 BMI values. A height recording was available for 20% of the visits, weight for 43%, head circumference for 6%, and BMI for 23% (supplementary Table S4). The availability of anthropometric recordings varied across age groups, types of visit, and between hospitals (Figure 1 and supplementary Figure S2). Height and weight were more often recorded during visits of infants and Swiss children compared to older and non-Swiss children (supplementary Tables S5 and S6). 38% of patients had at least one height, 76% at least one weight, and 14% at least one head circumference recording (supplementary Table 7). 38% of patients had a height recording within 30 days of a weight recording, to allow for calculation of BMI values. 23% of patients had multiple height and 49% multiple weight measurements, potentially allowing for longitudinal analyses.

**Figure 1.**
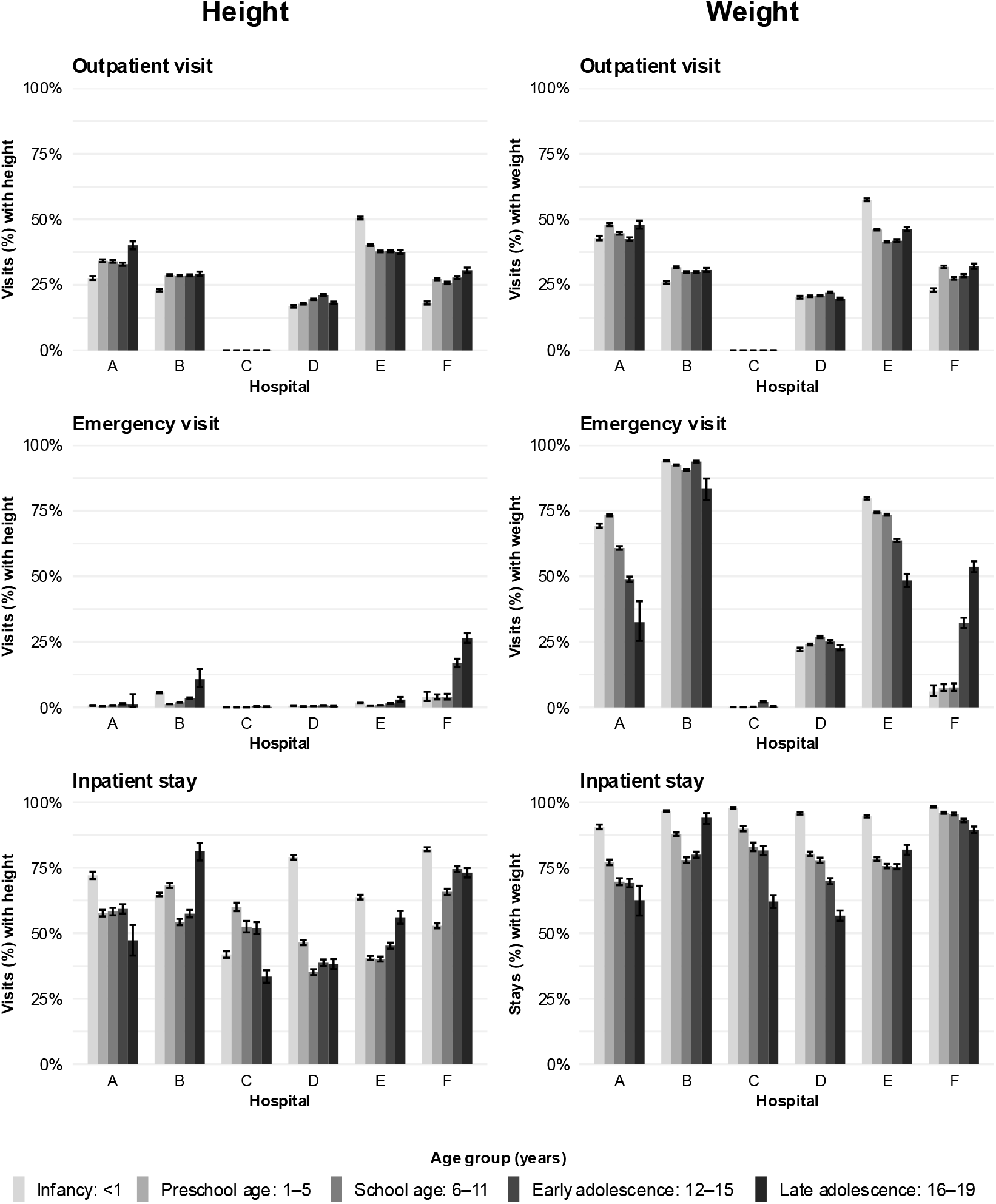
Proportion of hospital visits with an available height or weight recording in electronic health records of SwissPedGrowth hospitals stratified by type of visit, age group at visit, and hospital.

### 3.3 Quality of anthropometric data

Our self-developed algorithm flagged more biologically implausible outliers and duplicate recordings and corrected more errors than the growthcleanr (Figure 2 and supplementary Tables S8 and S9). Combining the two algorithms, we identified 159 (<1%) height and 184 (<1%) weight invalid recordings (zero or negative value, negative age), 28,553 (4%) height and 52,434 (3%) weight recordings as biologically implausible outliers, 195,359 (29%) heights and 533,651 (31%) weights as duplicate recordings, corrected 540 (<1%) errors of height (e.g. unit or decimal errors, recorded as weight) and 977 (<1%) errors of weight recordings, and found 451,054 (67%) height and 1,160,316 (66%) weight recordings without any error (Figure 2 and supplementary Table S10). Figure 3 illustrates the height and weight recordings plotted by age in the original dataset, during data cleaning, and in the cleaned dataset.

**Figure 2.**
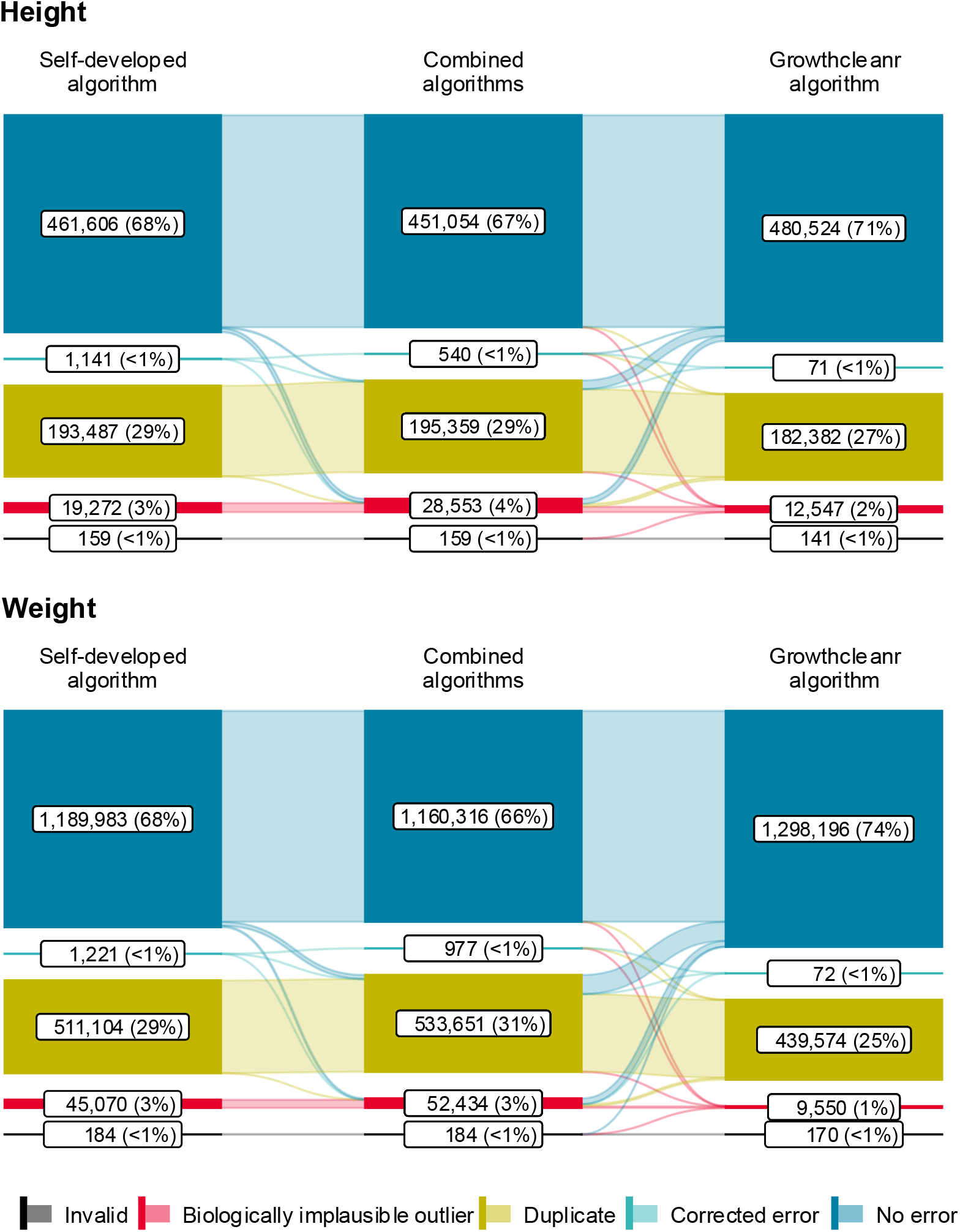
Quality of A) height and B) weight data extracted from electronic health records of SwissPedGrowth hospitals checked by our study-specific algorithm, the *growthcleanr* algorithm, and both algorithms combined. Our study-specific algorithm was based on z-scores of anthropometric measurements and the growthcleanr algorithm on a moving average of z-scores. See supplementary material for details about the algorithms.

**Figure 3.**
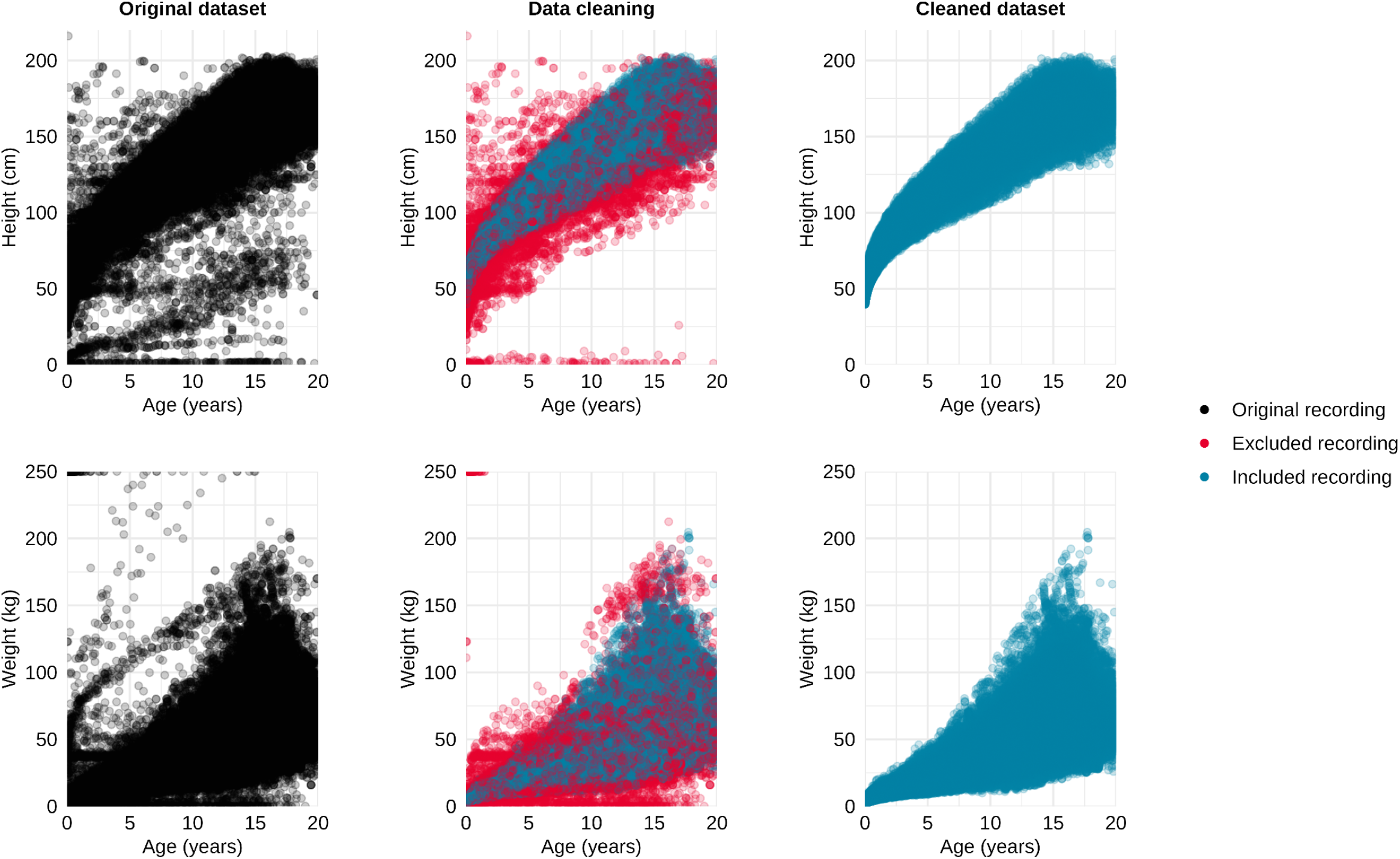
Included versus excluded height and weight recordings extracted from electronic health records of SwissPedGrowth hospitals. In this figure duplicates, biologically implausible outliers, and invalid recordings were excluded using our study-specific algorithm and the growthcleanr algorithm combined. Our study-specific algorithm was based on z-scores of anthropometric measurements and the growthcleanr algorithm on a moving average of z-scores. See supplementary material for details about the algorithms.

Our self-developed algorithm identified 167 (<1%) invalid head circumference recordings, 14,274 (7%) biologically implausible outliers, 34,685 (18%) duplicate recordings, and corrected 19 (<1%) decimal point errors (supplementary Table S11). For BMI, the combined algorithms identified 157 (<1%) invalid recordings, 66,547 (6%) biologically implausible outliers, 441,524 (38%) duplicated recordings, and 821 (<1%) corrected errors in the respective height or weight recordings of the calculated BMI values (supplementary Table S12). Additional 2,413 (<1%) calculated BMI values were flagged as biologically implausible outliers based on the BMI z-score. Supplementary Figure S3 illustrates the head circumference recordings and calculated BMI values in the original dataset, during data cleaning, and in the cleaned dataset.

### 3.4 Challenges of extracting electronic health records and their harmonization

We faced challenges extracting EHR data, harmonizing it across hospitals, and preparing them for analysis. We observed many missing values because EHRs often contained unstructured free text that was inaccessible to extract or the EHRs were less granular than the SPHN RDF schema. For example, diagnoses for outpatient visits were mostly missing because they were not captured in structured fields. The detailed SPHN RDF schema and use of external terminologies improved harmonization but created substantial workload at the CDWs of the hospitals. The conversion from the RDF graph to tabular data was computationally demanding. Additionally, the SPHN RDF schema linked clinical information not to healthcare encounters but only to administrative cases. The source systems of some hospitals did not carry forward links between variables and administrative cases, which complicated data mapping. For example, Hospital C had a systemic difficulty mapping height and weight recordings for outpatient and emergency visits (Figure 1). Administrative cases can include multiple outpatient visits happening weeks or months apart. A visit could include multiple healthcare encounters with timestamps on the same day or few days apart (e.g. blood draw, doctor consultation, finalization of documentation two days later). Hence, it was difficult to define outpatient visits and link variables to those visits. We tried to solve this using a combination of patient identifiers, case identifiers, and dates and timestamps.

The manual review of inpatient (n=70) and outpatient (n=70) visits without automatically extracted height and weight recordings showed the following: Nearly all of the reviewed inpatient visits had a weight recording (93%), but the recordings were often not captured in the dedicated structured field for weight. For example, weight was captured in scanned paper documents (e.g. premedication protocols of anaesthesia, nursing charts before introduction of EHRs). Height was found in 60% of reviewed inpatient stays, but was also often captured in unstructured fields. For the reviewed outpatient visits, weight was found in 56% and height in 35%, often captured in free text fields to create letters or reports (e.g. status or clinical summary fields).

### 3.5 Representativeness of SwissPedGrowth cohort relative to the general population

Compared to the general population of children <20 years living in Switzerland, the SwissPedGrowth population for the years 2017 and 2023 was younger, included more boys, fewer Swiss children, and more children of high socioeconomic status compared to the general population of children living in Switzerland (supplementary Table S13 and S14). The standardized differences in sex (Cohen’s d: 2017: 0.082, 2023: 0.072), nationality (Cohen’s d: 2017: 0.180, 2023: 0.098), and Swiss-SEP quintiles (Cohen’s d: 2017: 0.204, 2023: 0.220) were of small effect size, differences in age (Cohen’s d: 2017: 0.481, 2023: 0.371) of small-to-medium effect size. After weighting, the standardized differences were essentially zero (Cohen’s d for age, sex, nationality, and Swiss-SEP quintiles all <0.001).

## 4 DISCUSSION

### 4.1 Principal findings

Using EHRs remains challenging for paediatric growth research in Switzerland, primarily due to the substantial workload required to extract and harmonize data from heterogeneous EHR systems across hospitals. We retrieved less anthropometric data than expected: height recordings were found in 20% of the visits and weight in 43%. Only few height and weight recordings were biologically implausible outliers, but nearly one third were flagged as carried forward or same day duplicates. A manual review of 140 randomly selected visits revealed that much data from scanned documents or unstructured free text fields is missed. Notably, the SwissPedGrowth population was broadly representative of the Swiss general population regarding age, sex, and Swiss-SEP, especially after applying weighting.

### 4.2 Strengths and limitations

Strengths include the large sample size and multicentre design, rigorous efforts of harmonization of EHRs across children’s hospitals, and inclusion of patients without returned general consent. We included patients who were informed about general consent but had not returned the consent form (13%) and those with an unknown consent status (18%). By including this group, our study represents essentially all patients, except the 9% who actively refused general consent—similar to a previous Swiss study (10%) [29]— and reflects a real-world population.

Limitations include the lack of data the Italian speaking part of Switzerland, reducing the national representativeness. Furthermore, our reliance on structured fields likely led to lower data retrieval for outpatients and emergency visits compared with inpatients. We also had to exclude 15% of patients due to difficulties linking data to specific visits. Anthropometric parameters may be less rigorously measured in hospitals compared to studies focusing on childhood growth, and some recordings may be parent-reported. Finally, our self-developed cleaning algorithm may have overestimated biologically implausible outliers compared to established tools like the growthcleanr algorithm, as it was not validated against a manual paediatrician review of anthropometric recordings.

### 4.3 Comparison with other studies

The challenges of EHR extraction and harmonization are similar to those of large US networks like the Paediatric EHR Data Sharing Network (PEDSnet) or the San Diego EHR-Based Healthy Weight Surveillance System [7, 9]. PEDSnet showed that differences in data structures and coding of clinical data are barriers of multi-institutional EHR harmonization [7]. In France, new growth references were constructed using EHR data from primary care; harmonization was facilitated by including practices using the same EHR system [4]. Across Swiss children’s hospitals, harmonization was improved by the SwissPedHealth and SPHN frameworks [12], but unstructured data entry and heterogeneous EHR systems led to a huge workload to retrieve and make EHR data comparable across institutions.

Data availability in SwissPedGrowth was lower than in international counterparts. In PEDSnet, height was available in 58% of outpatient or well-child visits, weight in 69% [3]. Canadian primary care reached 68% for height and 98% for weight [30]. In contrast, we found height only in 27% of outpatient visits; weight in 31%. This discrepancy may be due to less freetext in PEDSnet and because they included well-child visits and outpatient visits, whereas we only included outpatient visits, which are shorter and usually not focused on a child’s development and growth.

Regarding data quality, our rate of detected biologically implausible outliers (1–2%) and same day recordings (7–11%) by the growthcleanr algorithm aligned with previous studies (1–2% outliers, 6–19% same day recordings) [5, 31, 32]. Our self-developed algorithm based on absolute z-scores, excluded more height (3%) and weight recordings (3%), than the French study extracting primary care data and also using absolute z-scores (0.3%) [4]. This suggests that capture of anthropometric data in hospitals may be more prone to errors than in primary care. More head circumference values were flagged as outliers (7%), suggesting head circumference is particularly vulnerable to measurement inaccuracies. SwissPedGrowth also had a higher proportion of carried forward values (14–20%) than reported previously (5–8%) [31, 32].

### 4.4 Implications

To improve the utility of EHRs for research, we suggest interventions at three levels: On the physician-level, training is required on the importance of entering clinical information in a structured manner, using less free text fields, to improve data completeness when EHRs are extracted. On the hospital-level, EHR systems must be optimized to facilitate structured data entry, especially also for high-volume outpatient and emergency visits. Improving algorithmic detection and extraction of data from these consultations could increase the amount of normal anthropometric information unaffected by acute or chronic diseases. Across hospitals, the SPHN framework should be adapted to more closely reflect actual EHR structures, which is difficult because of heterogeneous EHR systems. Specifically, providing clearer ways to link data to single visits rather than broad administrative cases would improve data interpretability and research readiness.

### 4.5 Conclusions

Despite the significant workload involved in multi-institutional harmonization, the SwissPedGrowth study demonstrates that the SwissPedHealth and SPHN framework make large-scale EHR research feasible. EHRs provide a large, high-quality anthropometric data source that, when properly weighted, can be representative of the general population. This should enable robust paediatric growth research answering both individual-level and public health questions.

## Data Availability

Data may be made available to investigators upon request by email to the corresponding author.

## GLOSSARY

EHR: Electronic health records
CDW: Clinical data warehouse
RDF: Resource description framework
SPHN: Swiss Personalized Health Network
SETT: Secure encryption and transfer tool
BMI: Body mass index

## AUTHOR CONTRIBUTIONS

CEK, JAB, PL, KMPB, EG, MS, RL, RM, FB, BS, and LJS conceptualized the study and acquired funding. FB and RM set up and coordinated the study. XD, FB, RM, AH, YS, and LML contributed to data curation. LML conducted the formal analysis with support from YS, FR, and BS under the supervision of FB and CEK. MAB, CS, VS, FRG, MH, PM, and OGJ conducted a manual review at their hospital to validate the data. All authors interpreted and discussed the findings. LML wrote the original draft, all authors critically reviewed and edited the manuscript. All authors approved the final version of the manuscript.

## DATA STATEMENT

Data may be made available to investigators upon request by email to the corresponding author.

