## Supplementary Material for "Availability and Quality of Anthropometric Data in Swiss Children’s Hospitals: The SwissPedGrowth Project"

### Supplementary Methods

#### Data extraction and preparation

Hospitals participating in the SwissPedHealth national data stream extracted sociodemographic, administrative, anthropometric, and clinical data from electronic capture systems and other source systems and loaded it into local clinical data warehouses (CDWs) [1]. They linked the Neighbourhood index of socioeconomic position (Swiss-SEP) to the patient’s home address using a standard operating procedure [2, 3]. They mapped the data to the schema of the Swiss Personalized Health Network (SPHN) that is structured into so called concepts (variable group, e.g. AdministrativeCase) and composedOf variables (metadata for this variable group, e.g. Admission or Discharge of an AdministrativeCase) that link to standard terminologies, including ATC, LOINC, SNOMED CT, UCUM, and more [4]. Using the SPHN Connector the data was transformed into the Resource Description Format (RDF)—a graph based data format [5]. CDWHs encrypted the data and sent it via the Secure Encryption and Transfer Tool (sett) to BioMedIT—a secure server for processing and analysing medical data. Data managers of SwissPedHealth filtered the required variables for SwissPedGrowth from the RDF database of SwissPedHealth and transferred the filtered data to the SwissPedGrowth project on BioMedIT.

We then used SPARQL queries (SPARQL Protocol and RDF Query Language) in the GraphDB software to transform each concept and its composedOf variables from the RDF data into a comma separated value (csv) tabular data format. We imported the csv files into the statistical software R and coded the variables needed for analysis, see below. We merged patient characteristic concepts together into a dataset of patients using the patient identifier. We merged administrative concepts (AdministrativeCase and HealthcareEncounter) together into a dataset of hospital visits using patient identifiers, case identifiers and dates.

Hospitals coded CareHandling information of an AdministrativeCase as “Outpatient procedure” or “Provision of day care” if care was provided ambulatory and as “Inpatient care” if care included an overnight stay. We defined three types of visits based on information in the AdministrativeCase and HealthcareEncounter concepts:

i) **Outpatient visit:** HealthcareEncounters with “Outpatient procedure” or “Provision of day care” as CareHandling information. We combined all outpatient HealthcareEncounters of the same patient, the same AdministrativeCase, and the same week into one single outpatient visit.
ii) **Emergency visit:** HealthcareEncounters that happened at an emergency department. We combined all emergency HealthcareEncounters of the same patient, the same AdministrativeCase, and the same week into one single emergency visit.
iii) **Inpatient stay:** HealthcareEncounters with “Inpatient care” as CareHandling information. We combined all inpatient HealthcareEncounters of the same patient and the same AdminisitrativeCase into one single inpatient stay.

To create a dataset of body mass index (BMI), we took weight recordings and matched the closest height recording allowing for a maximum of 30 days between the weight and height recording. We created BMI after assessing the quality of height and weight recordings and correcting errors to prioritize good quality recordings to get matched. We calculated BMI as weight[kg]/height[cm]^2^. Finally, we merged the patient and visit datasets to the diagnosis, height, weight, head circumference, and BMI datasets using patient identifiers, case identifiers, and dates. The datasets, SPHN concepts, and SPHN composedOf are described in supplementary table 1.

#### Linking the Swiss-SEP to patients’ addresses

The Neighbourhood index of socioeconomic position (Swiss-SEP) was created by the Swiss National Cohort (SNC) [6]. The Swiss-SEP is an area-based measure of socioeconomic status and was calculated for 1.27 million overlapping neighbourhoods using information from the Swiss census of the year 2000, and updated for 1.54 million overlapping neighbourhoods with information from the 2012–2015 yearly micro censuses [2, 6]. We linked the Swiss-SEP to patients’ addresses in two steps. The standard operating procedure (SOP) is published on the Bern Open Repository and Information System (BORIS) [3]; the code is available on [GitHub](https://github.com/LorenzLeuenberger/Transforming-addresses-to-Swiss-SEP_Standard-operating-procedure).

i) **Link Swiss-SEP to all addresses in Switzerland:** We downloaded the Swiss federal building register containing addresses and geographic coordinates of all buildings in Switzerland. The addresses were already tokenized (street, number, postal code, city) and additionally contained abbreviated street names. We spelled out umlauts and removed accents or special characters from the addresses and created an additional numerical token for the number. The Swiss-SEP dataset was shared by the SNC after signing a data delivery contract, containing Swiss-SEP values and geographic coordinates of the neighbourhood. We linked the geographically closest Swiss-SEP value to every address, using the *sf* package in R. This resulted in a reference dataset of all Swiss addresses and linked Swiss-SEP values. We prepared this reference dataset centrally and distributed it to the CDWHs of the hospitals.
ii) **Look up patients’ addresses:** The CDWHs used the SOP to look up the patients’ addresses in the reference dataset. We first split patients’ addresses into four tokens: street, number, postal code, and city. We spelled out common German, French, and Italian abbreviations for street names, spelled out umlauts, and removed special characters. We also created the numerical token for the number. We then used two steps for looking up the patient addresses in the reference dataset.
    a. First, we looked up addresses with exact matching street, number, postal code, and city. We used multiple rounds to exactly match addresses using the different tokens for the street (spelled out or abbreviated in the reference dataset), number (character or numeric), and city (postal code or city).
    b. Second, we looked up non-matched patients’ addresses using custom functions to fuzzy match them to addresses in the reference dataset. We calculated a matching score for street, number, postal code, and city tokens using numerical rules for numeric tokens and Levenstein distance for character tokens. The matching score could reach a maximum “non-match” if the score of a single token or the combined score of all tokens became too high. To reduce computations, we limited comparisons to addresses in the reference dataset that had the same postal code or city as the patient’s address. We then chose the reference address with the lowest matching score. We coded this SOP in R and python and translated it to SQL.

#### Algorithms to check data quality of anthropometric measurements

To assess data quality, we identified unit errors, decimal errors, swapped recordings (height recorded as weight and vice versa), duplicated recordings, biologically implausible outliers, and invalid recordings using three stages: a self-developed algorithm, the existing growthcleanr [3] algorithm, and both algorithms combined.

i) **For our self-developed algorithm**, we calculated height, weight, and head circumference z-scores based on the WHO growth references adopted to Switzerland [7]. We flagged recordings as “invalid” if they had a zero or negative value or a negative age at recording. To identify swapped recordings, we interpreted height recordings as weight recordings and vice versa if they had an extreme z-score <-10 or >10 and recalculated the z-scores. If the new z-score was closer to zero than the original z-score and >-5 and <5 for height or >-5 and <8 for weight, we flagged the recordings as “swapped recording” and corrected them. To identify unit errors, we converted recordings to a different unit (e.g. 1.05 cm to 1.05 m) if they had an extreme z-score <-10 or >10 and recalculated the z-scores. If the new z-score was closer to zero than the original z-score and >-5 and <5 for height and head circumference, or >-5 and <8 for weight, we flagged the recordings as “unit errors” and corrected them. To identify decimal point errors, we converted recordings to another decimal point (e.g. 10.5 cm to 105 cm) if they had an extreme z-score <-10 or >10, recalculated the z-scores, flagged them as “decimal point errors” if the above z-score criteria were met, and corrected them. To identify duplicated recordings, we searched for identical recordings of the same patient and flagged them as “carried forward” if they were not the first, because it is likely that the following recordings were not measured but copied from previous visits. For multiple recordings on the same date, we chose the one with the z-score closest to zero and flagged the others as “same day duplicates”. To identify biologically implausible values, we flagged height and head circumference recordings with a z-score <-5 and >5 and weight recordings <-5 and >8 as “z-score outlier”. We flagged recordings without any of the above errors as “no error”.
ii) **For the growthcleanr algorithm**, we used the cleangrowth function from the *growthcleanr* package in R on a combined dataset with height and weight recordings [8]. The only setting differing from default was to use the WHO growth references adopted to Switzerland [7]. We grouped the output of the growthcleanr into: “no error”, “unit error”, “swapped recordings” (swapped height and weight), “carried forward”, “same day”, “single outlier”, “longitudinal outlier”, and “invalid”.
iii) **For both algorithms combined**, we first ran our self-developed algorithm creating the flags and correcting swapped recordings, unit errors, and decimal point errors. Then, we ran the growthcleanr algorithm. We flagged the height and weight recordings combining the flags of both algorithms from “invalid information” if one of both algorithms flagged a recording as “invalid” up to “no error” if both algorithms flagged a recording as “no error”.

For head circumference recordings, we only used our self-developed algorithm.

For calculated BMI values, we used the results from both algorithms combined of the respective weight and height recordings and additionally flagged biologically implausible BMI values as z-scores <-5 or >8.

### Supplementary Figures and Tables

**Data in SwissPedGrowth**
Patients, N = 560,955

Visits, N = 2,346,460

Height recordings, N = 689,072
Weight recordings, N = 1,952,751
Head circumference recordings, N = 193,091
Calculated BMI, N = 1,193,046

**Excluded visits**

Missing sex, n = 1,007 (<1%)

Missing type of visit, n = 173,820 (7%)

**Excluded patients**

Missing sex, n = 83 (<1%)

Missing type of visit, n = 83,341 (15%)

**Data included in analysis**

Patients, N = 477,531 (85%)
Visits, N = 2,171,633 (93%)

Height recordings, N = 675,665 (98%)
Weight recordings, N = 1,747,562 (89%)
Head circumference recordings, N = 192,631 (>99%)
Calculated BMI, N = 1,169,075 (98%)

**Figure S1: Flow diagram of patients and visits extracted from electronic health records of SwissPedGrowth hospitals included in the analysis.**
We excluded patients with missing sex and visits for which we were unable to define the type of visit, e.g. because of missing information about the department of visit. We had to exclude all patients from St. Gallen because we did not receive birth dates and were unable to check the inclusion criteria of the SwissPedGrowth project; these patients and their data are therefore not shown in this flowchart.

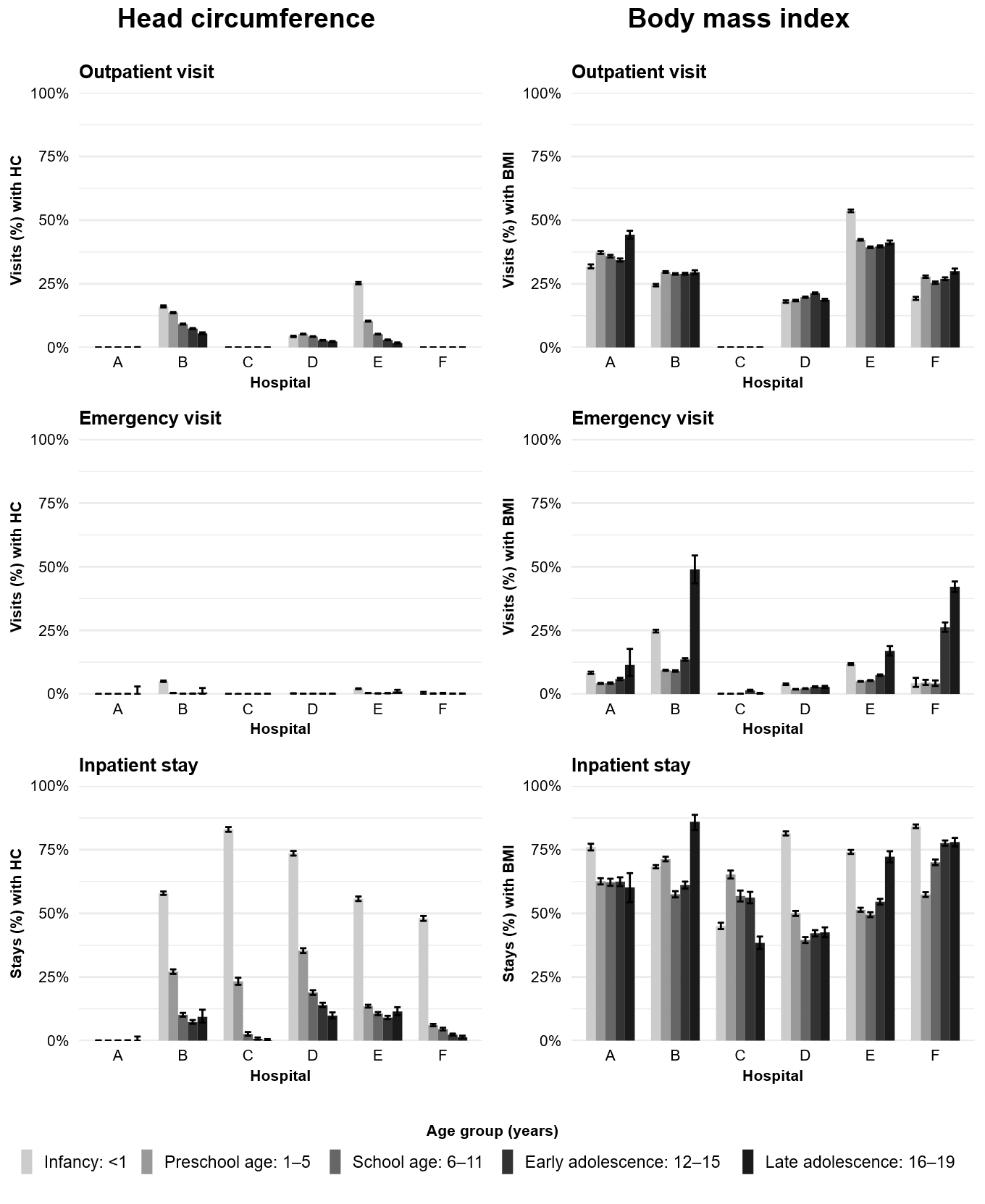

**Figure S2. Proportion of hospital visits with an available head circumference recording or a calculated body mass index value in electronic health records of SwissPedGrowth hospitals stratified by type of visit, age group at visit, and hospital.**

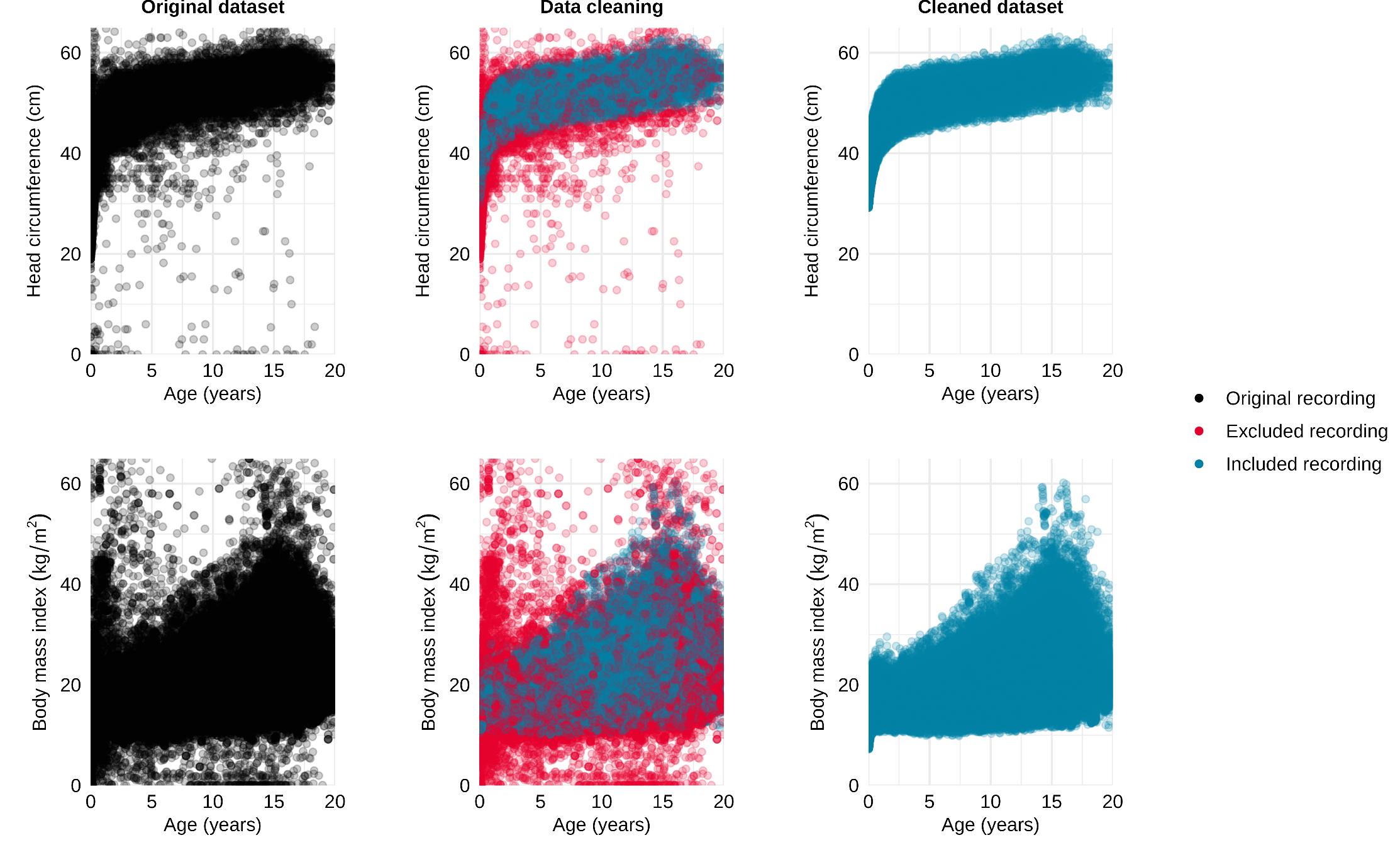

**Figure S3.** **Included versus excluded head circumference recordings extracted from electronic health records of SwissPedGrowth hospitals and calculated BMI values.**
In this figure duplicates, biologically implausible outliers, and invalid recordings were excluded. For head circumference we only used our self-developed algorithm. For BMI we used our self-developed algorithm and the growthcleanr algorithm combined to flag height and weight recordings; we additionally excluded BMI values with z-scores <-5 or >8. See supplementary material for details about the algorithms.

**Table S1. Description of datasets, SPHN concepts, and SPHN composedOf used in SwissPedGrowth.**

|  | **Variable** | **SPHN concept** | | | | | | **SPHN composedOf** | | | | **Data type** | | | | | | | **Description** | | | | | |
| --- | --- | --- | --- | --- | --- | --- | --- | --- | --- | --- | --- | --- | --- | --- | --- | --- | --- | --- | --- | --- | --- | --- | --- | --- |
| Patient dataset (linked by participant_id) | participant_id | SubjectPseudoIdentifier | | | | | | hasIdentifier | | | | xsd:string | | | | | | | Identifier to link SPHN concepts to an individual. | | | | | |
|  | hospital | SubjectPseudoIdentifier | | | | | | hasDataProvider/hasInstitutionCode | | | | UID | | | | | | | Universal institution code of hospital that provided the data.  Coded as: Basel, Bern, Geneva, Lausanne, Luzern, St. Gallen, Zurich | | | | | |
|  | birth_date | BirthDate | | | | | | hasYear | | | | xsd:gYear | | | | | | | Date and time of birth of an individual.  Coded as datetime. | | | | | |
|  |  | BirthDate | | | | | | hasMonth | | | | xsd:gMonth | | | | | | |  |  |  |  |  |  |
|  |  | BirthDate | | | | | | hasDay | | | | xsd:gDay | | | | | | |  |  |  |  |  |  |
|  |  | BirthDate | | | | | | hasTime | | | | xsd:time | | | | | | |  |  |  |  |  |  |
|  | participant_id | BirthDate | | | | | | hasSubjectPseudoIdentifier/hasIdentifier | | | | xsd:string | | | | | | | Identifier to link BirthDate to an individual. | | | | | |
|  | sex | AdministrativeSex | | | | | | hasCode | | | | SNOMED CT | | | | | | | Sex of an individual. Coded as: Female, Male | | | | | |
|  | participant_id | AdministrativeSex | | | | | | hasSubjectPseudoIdentifier/hasIdentifier | | | | xsd:string | | | | | | | Identifier to link AdministrativeSex to an individual. | | | | | |
|  | nationality | Nationality | | | | | | hasAssociatedCountry/hasCode | | | | SNOMED CT | | | | | | | Nationality of an individual. Coded as: Swiss, European, Non-European  See eTable 1 for a list of detailed nationalities. | | | | | |
|  | participant_id | Nationality | | | | | | hasSubjectPseudoIdentifier/hasIdentifier | | | | xsd:string | | | | | | | Identifier to link Nationality to an individual. | | | | | |
|  | swiss_sep_value | SwissSocioEconomicPosition | | | | | | hasValue | | | | xsd:double | | | | | | | Swiss neighbourhood index of socioeconomic position (Swiss-SEP), see Panczak et al. [2] Continuous (0–100) | | | | | |
|  | swiss_sep_version | SwissSocioEconomicPosition | | | | | | hasVersion | | | | xsd:string | | | | | | | Version of the Swiss-SEP, see Panczak et al. | | | | | |
|  | swiss_sep_3 | Coded from SwissSocioEconomicPosition/hasValue and hasVersion | | | | | | | | | | | | | | | | | Version 3.0 of the Swiss-SEP.  Continuous (0–100) | | | | | |
|  | swiss_sep_3_quintile | Coded from SwissSocioEconomicPosition/hasValue and hasVersion | | | | | | | | | | | | | | | | | Quintile of the Swiss-SEP version 3.0 based on the deciles published by Panczak et al.  Coded as quintiles: 1, 2, 3, 4, 5 | | | | | |
|  | participant_id | SwissSocioEconomicPosition | | | | | | hasSubjectPseudoIdentifier/hasIdentifier | | | | xsd:string | | | | | | | Identifier to link BirthDate to an individual. | | | | | |
|  | consent_type | Consent | | | | | | hasTypeCode | | | | SNOMED CT | | | | | | | Type of consent. For this analysis we used general hospital consent. | | | | | |
|  | consent_status | Consent | | | | | | hasStatusCode | | | | SNOMED CT | | | | | | | Status of the consent. Hospitals did not send any data of patients with declined general hospital consent.  Coded as: Accepted, Informed, Unknown | | | | | |
|  | participant_id | Consent | | | | | | hasSubjectPseudoIdentifier/hasIdentifier | | | | xsd:string | | | | | | | Identifier to link BirthDate to an individual. | | | | | |
|  | **Variable** | | **SPHN concept** | | | | **SPHN composedOf** | | | | | | **Data type** | | | | | | | **Description** | | | | |
| Visit dataset (linked by case_id) | case_id | | AdministrativeCase | | | | hasIdentifier | | | | | | xsd:string | | | | | | | Administrative artefact used for billing.  Identifier to link SPHN concepts to an administrative case. | | | | |
|  | participant_id | | AdministrativeCase | | | | hasSubjectPseudoIdentifier/hasIdentifier | | | | | | xsd:string | | | | | | | Identifier to link AdministrativeCase to an individual. | | | | |
|  | case_admission_datetime | | AdministrativeCase | | | | hasAdmission/hasDateTime | | | | | | xsd:dateTime | | | | | | | Date and time of admission of a patient to the hospital. | | | | |
|  | case_discharge_datetime | | AdministrativeCase | | | | hasDischarge/hasDateTime | | | | | | xsd:dateTime | | | | | | | Date and time of discharge of a patient to the hospital. | | | | |
|  | case_duration | | Coded from case_admission_datetime and case_discharge_datetime | | | | | | | | | | | | | | | | | Duration of administrative case.  Coded separately as hours and as days. | | | | |
|  | age_case_admission | | Coded from birth_date and case_admission_datetime | | | | | | | | | | | | | | | | | Age of patient at admission.  Coded separately as days and as years. | | | | |
|  | age_case_admission | | Coded from birth_date and case_discharge_datetime | | | | | | | | | | | | | | | | | Age of patient at discharge.  Coded separately as days and as years. | | | | |
|  | case_carehandling | | AdministrativeCase | | | | hasCareHandling | | | | | | SNOMED CT | | | | | | | Relationship between the individual and the hospital.  Coded as: Inpatient care, Outpatient procedure, Provision of day care | | | | |
|  | encounter_id | | HealthcareEncounter | | | | hasIdentifier | | | | | | xsd:string | | | | | | | Interaction between an individual and a specific unit or service of a hospital. | | | | |
|  | participant_id | | HealthcareEncounter | | | | hasSubjectPseudoIdentifier/hasIdentifier | | | | | | xsd:string | | | | | | | Identifier to link HealthcareEncounter to an individual. | | | | |
|  | case_id | | HealthcareEncounter | | | | hasAdministrativeCase/hasIdentifier | | | | | | xsd:string | | | | | | | Identifier to link HealthcareEncounter to an administrative case. | | | | |
|  | encounter_current_location_code | | HealthcareEncounter | | | | hasCurrentLocation/hasTypeCode | | | | | | SNOMED CT | | | | | | | Department where the visit happened (e.g. »Pediatric Accident and Emergency department«, »Hospital-based outpatient pediatric clinic«) | | | | |
|  | encounter_start_datetime | | HealthcareEncounter | | | | hasStartDateTime | | | | | | xsd:dateTime | | | | | | | Date and time when the encounter started. | | | | |
|  | encounter_end_datetime | | HealthcareEncounter | | | | hasEndDateTime | | | | | | xsd:dateTime | | | | | | | Date and time when the encounter ended. | | | | |
|  | encounter_duration | | Coded from encounter_start_datetime and encounter_end_datetime | | | | | | | | | | | | | | | | | Duration of encounter.  Coded separately as minutes, as hours, and as days. | | | | |
|  | age_encounter_start | | Coded from birth_date and encounter_start_datetime | | | | | | | | | | | | | | | | | Age of patient at encounter start.  Coded separately as days and years. | | | | |
|  | age_encounter_end | | Coded from birth_date and encounter_end_datetime | | | | | | | | | | | | | | | | | Age of patient at encounter end.  Coded separately as days and years. | | | | |
|  | encounter_carehandling | | Coded from case_carehandling and encounter_current_location_code | | | | | | | | | | | | | | | | | Type of encounter.  Coded as: Inpatient encounter, outpatient encounter, emergency encounter | | | | |
|  | visit_id | | Coded from case_carehandling and encounter_current_location_code | | | | | | | | | | | | | | | | | Hospital visits defined by us:   i) Outpatient visit: HealthcareEncounters with “Outpatient procedure” or “Provision of day care” as CareHandling information. We combined all outpatient HealthcareEncounters of the same patient, the same AdministrativeCase, and the same week into one single outpatient visit. ii) Emergency visit: HealthcareEncounters that happened at an emergency department. We combined all emergency HealthcareEncounters of the same patient, the same AdministrativeCase, and the same week into one single emergency visit. iii) Inpatient stay: HealthcareEncounters with “Inpatient care” as CareHandling information. We combined all inpatient HealthcareEncounters of the same patient and the same AdminisitrativeCase into one single inpatient stay. | | | | |
|  | visit_start_datetime | | Coded from case_admission_datetime or encounter_start_datetime | | | | | | | | | | | | | | | | | Date and time when the hospital visit started.  AdministrativeCase/hasAdmission for inpatient visits. First HealthcareEncounter/hasStartDateTime for outpatient visits and emergency visits. | | | | |
|  | visit_end_datetime | | Coded from case_discharge_datetime or encounter_end_datetime | | | | | | | | | | | | | | | | | Date and time when the hospital visit ended.  AdministrativeCase/hasDischarge for inpatient visits. Last HealthcareEncounter/hasEndDateTime for outpatient visits and emergency visits. | | | | |
|  | age_visit_start | | Coded from birth_date and visit_start_datetime | | | | | | | | | | | | | | | | | Age of patient at start of hospital visit.  Coded as years. | | | | |
|  | age_visit_end | | Coded from birth_date and visit_end_datetime | | | | | | | | | | | | | | | | | Age of patient at end of hospital visit.  Coded as years. | | | | |
|  | visit_type | | Coded from AdministrativeCase/hasCarehandling and HealthcareEncounter/hasLocation | | | | | | | | | | | | | | | | | Type of visit.  Coded as: inpatient stay, outpatient visit, emergency visit | | | | |
|  | **Variable** | | | | | **SPHN concept** | **SPHN composedOf** | | | | | | | **Data type** | | | | | | | **Description** | | | |
| Diagnosis dataset | diagnosis_id | | | | | BilledDiagnosis |  | | | | | | | xsd:string | | | | | | | Discharge diagnosis used for the billing system. | | | |
|  | participant_id | | | | | BilledDiagnosis | hasSubjectPseudoIdentifier/hasIdentifier | | | | | | | xsd:string | | | | | | | Identifier to link BilledDiagnosis to an individual. | | | |
|  | case_id | | | | | BilledDiagnosis | hasAdministrativeCase/hasIdentifier | | | | | | | xsd:string | | | | | | | Identifier to link BilledDiagnosis to an administrative case. | | | |
|  | diagnosis_icd10_code | | | | | BilledDiagnosis | hasCode | | | | | | | ICD-10 GM | | | | | | | Diagnosis code in the International Classification of Diseases 10^th^ revision, german modification | | | |
|  | diagnosis_icd10_code_label | | | | | Extracted from BilledDiagnosis/hasCode and the associated rdfs:label | | | | | | | | | | | | | | | Label of the ICD-10 diagnosis code. | | | |
|  | diagnosis_record_datetime | | | | | BilledDiagnosis | hasRecordDateTime | | | | | | | xsd:dateTime | | | | | | | Time of recording of the diagnosis. | | | |
|  | age_diagnosis | | | | | Coded from birth_date and diagnosis_record_datetime or from BilledDiagnosis/hasSubjectAge | | | | | | | | | | | | | | | Age of patient at time of diagnosis.  Coded separately as days and years. | | | |
|  | diagnosis_id | | | | | Diagnosis |  | | | | | | | xsd:string | | | | | | | Discharge diagnosis used for the billing system. | | | |
|  | participant_id | | | | | Diagnosis | hasSubjectPseudoIdentifier/hasIdentifier | | | | | | | xsd:string | | | | | | | Identifier to link Diagnosis to an individual. | | | |
|  | case_id | | | | | Diagnosis | hasAdministrativeCase/hasIdentifier | | | | | | | xsd:string | | | | | | | Identifier to link Diagnosis to an administrative case. | | | |
|  | diagnosis_icd10_code | | | | | Diagnosis | hasCode | | | | | | | ICD-10 GM | | | | | | | Diagnosis code in the International Classification of Diseases 10^th^ revision, german modification | | | |
|  | diagnosis_icd10_code_label | | | | | Extracted from Diagnosis/hasCode and the associated rdfs:label | | | | | | | | | | | | | | | Label of the ICD-10 diagnosis code. | | | |
|  | diagnosis_orpha_code | | | | | Diagnosis | hasCode | | | | | | | ORPHAcode | | | | | | | Diagnosis code in the nomenclature of rare diseases, orphanet | | | |
|  | diagnosis_orpha_code_label | | | | | Extracted from Diagnosis/hasCode and the associated rdfs:label | | | | | | | | | | | | | | | Label of the Orphanet diagnosis code. | | | |
|  | diagnosis_orpha_code_associated_icd | | | | | Extracted from Diagnosis/hasCode and the associated oboInOwl:hasDbXref | | | | | | | | | | | | | | | ICD-10 diagnosis code that is associated to the Orphanet diagnosis code. | | | |
|  | diagnosis_record_datetime | | | | | Diagnosis | hasRecordDateTime | | | | xsd:dateTime | | | | | | | | | | Time of recording of the diagnosis. | | | |
|  | age_diagnosis | | | | | Coded from birth_date and diagnosis_record_datetime or from Diagnosis/hasSubjectAge | | | | | | | | | | | | | | | Age of patient at time of diagnosis.  Coded separately as days and years. | | | |
|  | diagnosis_affecting_growth | | | | | Coded from diagnosis_icd10_code and diagnosis_orpha_code_associated_icd | | | | | | | | | | | | | | | Expert opinion whether diagnosis affects growth, see appendix classification of ICD-10 codes.  Coded as: affecting growth permanently, affecting growth temporarily, not affecting growth | | | |
|  | **Variable** | | | | **SPHN concept** | | | | | **SPHN composedOf** | | | | | | **Data type** | | | | | | **Description** | | |
| Height dataset | height_id | | | | BodyHeightMeasurement | | | | |  | | | | | | xsd:string | | | | | | Recording of the height of an individual. | | |
|  | participant_id | | | | BodyHeightMeasurement | | | | | hasSubjectPseudoIdentifier/hasIdentifier | | | | | | xsd:string | | | | | | Identifier to link BodyHeightMeasurement to an individual. | | |
|  | case_id | | | | BodyHeightMeasurement | | | | | hasAdministrativeCase/hasIdentifier | | | | | | xsd:string | | | | | | Identifier to link BodyHeightMeasurement to an administrative case. | | |
|  | height_value | | | | BodyHeightMeasurement | | | | | hasBodyHeight/hasQuantity/hasValue | | | | | | xsd:double | | | | | | Value of the height recording. | | |
|  | height_unit | | | | BodyHeightMeasurement | | | | | hasBodyHeight/hasQuantity/hasUnit/hasCode | | | | | | UCUM | | | | | | Unit of the height recording in the Unified Code for Units of Measure (UCUM). | | |
|  | height_cm | | | | Coded from hasValue and hasUnit | | | | | | | | | | | | | | | | | Height recording in centimeters. | | |
|  | height_datetime | | | | BodyHeightMeasurement | | | | | hasStartDateTime  or hasBodyHeight/hasDateTime | | | | | xsd:dateTime | | | | | | | Date and time of the height recording. | | |
|  | age_height | | | | Coded from birth_date and height_datetime | | | | | | | | | | | | | | | | | Age of patient at time of height recording.  Coded separately as days and years. | | |
|  | z_height_braegger | | | | Coded from height_cm, age_height, sex, and LMS-Parameters of the WHO growth charts | | | | | | | | | | | | | | | | | Age- and sex-specific z-score of the height recording based on the WHO growth charts adopted to Switzerland [7]. | | |
|  | **Variable** | | | | **SPHN concept** | | | | | **SPHN composedOf** | | | | | | | | **Data type** | | | | | **Description** | |
| Weight dataset | weight_id | | | | BodyWeightMeasurement | | | | |  | | | | | | | | xsd:string | | | | | Recording of the weight of an individual. | |
|  | participant_id | | | | BodyWeightMeasurement | | | | | hasSubjectPseudoIdentifier/hasIdentifier | | | | | | | | xsd:string | | | | | Identifier to link BodyWeightMeasurement to an individual. | |
|  | case_id | | | | BodyWeightMeasurement | | | | | hasAdministrativeCase/hasIdentifier | | | | | | | | xsd:string | | | | | Identifier to link BodyWeightMeasurement to an administrative case. | |
|  | weight_value | | | | BodyWeightMeasurement | | | | | hasBodyWeight/hasQuantity/hasValue | | | | | | | | xsd:double | | | | | Value of the weight recording. | |
|  | weight_unit | | | | BodyWeightMeasurement | | | | | hasBodyWeight/hasQuantity/hasUnit/hasCode | | | | | | | | UCUM | | | | | Unit of the weight recording in the Unified Code for Units of Measure (UCUM). | |
|  | weight_kg | | | | Coded from hasValue and hasUnit | | | | | | | | | | | | | | | | | | Weight recording in kilograms. | |
|  | weight_datetime | | | | BodyWeightMeasurement | | | | | hasStartDateTime or hasBodyWeight/hasDateTime | | | | | | | | xsd:dateTime | | | | | Date and time of the weight recording. | |
|  | age_weight | | | | Coded from birth_date and weight_datetime | | | | | | | | | | | | | | | | | | Age of patient at time of weight recording.  Coded separately as days and years. | |
|  | z_weight_braegger | | | | Coded from weight_kg, age_weight, sex, and LMS-Parameters of the WHO growth charts | | | | | | | | | | | | | | | | | | Age- and sex-specific z-score of the weight recording based on the WHO growth charts adopted to Switzerland [7]. | |
|  | **Variable** | | | | **SPHN concept** | | | | | **SPHN composedOf** | | | | | | | | **Data type** | | | | | **Description** | |
| Head circumference dataset | circumference_id | | | | CircumferenceMeasurement | | | | |  | | | | | | | | xsd:string | | | | | Recording of the circumference of a body site of an individual. | |
|  | participant_id | | | | CircumferenceMeasurement | | | | | hasSubjectPseudoIdentifier/hasIdentifier | | | | | | | | xsd:string | | | | | Identifier to link CircumferenceMeasurement to an individual. | |
|  | case_id | | | | CircumferenceMeasurement | | | | | hasAdministrativeCase/hasIdentifier | | | | | | | | xsd:string | | | | | Identifier to link CircumferenceMeasurement to an administrative case. | |
|  | circumference_value | | | | CircumferenceMeasurement | | | | | hasBodyHeight/hasQuantity/hasValue | | | | | | | | xsd:double | | | | | Value of the circumference recording. | |
|  | circumference_unit | | | | CircumferenceMeasurement | | | | | hasBodyHeight/hasQuantity/hasUnit/hasCode | | | | | | | | UCUM | | | | | Unit of the circumference recording in the Unified Code for Units of Measure (UCUM). | |
|  | circumference_bodysite | | | | CircumferenceMeasurement | | | | | hasBodySite/hasCode | | | | | | | | SNOMED CT | | | | | Body site of the circumference recording. For this dataset we selected circumference recordings of the head (»Head structure«). | |
|  | head_cm | | | | Coded from hasValue and hasUnit | | | | | | | | | | | | | | | | | | Head circumference recording in centimeters. | |
|  | circumference_datetime | | | | CircumferenceMeasurement | | | | | hasStartDateTime or hasCircumference/hasDateTime | | | | | | | | xsd:dateTime | | | | | Date and time of the circumference recording. | |
|  | age_circumference | | | | Coded from birth_date and circumference_datetime | | | | | | | | | | | | | | | | | | Age of patient at time of circumference recording.  Coded separately as days and years. | |
|  | z_head_braegger | | | | Coded from head_cm, age_height, sex, and LMS-Parameters of the WHO growth charts adopted to Switzerland by Braegger et al. [7] | | | | | | | | | | | | | | | | | | Age- and sex-specific z-score of head circumference recording. | |
|  | **Variable** | | | **SPHN concept** | | | | | **SPHN composedOf** | | | | | | | | **Data type** | | | | | | | **Description** |
| Weight dataset | weight_id | | | BodyWeightMeasurement | | | | |  | | | | | | | | xsd:string | | | | | | | Recording of the weight of an individual. |
|  | participant_id | | | BodyWeightMeasurement | | | | | hasSubjectPseudoIdentifier/hasIdentifier | | | | | | | | xsd:string | | | | | | | Identifier to link BodyWeightMeasurement to an individual. |
|  | case_id | | | BodyWeightMeasurement | | | | | hasAdministrativeCase/hasIdentifier | | | | | | | | xsd:string | | | | | | | Identifier to link BodyWeightMeasurement to an administrative case. |
|  | weight_kg | | | Coded from hasValue and hasUnit | | | | | | | | | | | | | | | | | | | | Weight recording in kilograms. |
|  | weight_datetime | | | BodyWeightMeasurement | | | | | hasStartDateTime or hasBodyWeight/hasDateTime | | | | | | | | xsd:dateTime | | | | | | | Date and time of the weight recording. |
|  |  | | | BodyHeightMeasurement | | | | |  | | | | | | | | xsd:string | | | | | | | Recording of the height of an individual.  We linked the closest height recording within 30 days to a weight recording, prioritizing “included” height recordings first. |
|  | height_cm | | | Coded from hasValue and hasUnit | | | | | | | | | | | | | | | | | | | | Height recording in centimeters. |
|  | height_datetime | | | BodyHeightMeasurement | | | | | hasStartDateTime or hasBodyHeight/hasDateTime | | | | | | | | xsd:dateTime | | | | | | | Date and time of the height recording. |
|  | bmi | | | Coded from height_cm and weight_kg | | | | | | | | | | | | | | | | | | | | Calculated body mass index value of an individual. |
|  | age_bmi | | | Coded from birth_date and weight_datetime | | | | | | | | | | | | | | | | | | | | Age of patient at time of calculated BMI value, taken from age at weight recording.  Coded separately as days and years. |
|  | z_bmi_braegger | | | Coded from bmi, age_bmi, sex, and LMS-Parameters of the WHO growth charts | | | | | | | | | | | | | | | | | | | | Age- and sex-specific z-score of the weight recording based on the WHO growth charts adopted to Switzerland [7]. |

**Table S2. List of ICD-10 diagnosis codes and classification of affecting growth parameters**.

| **ICD-10 code** | **Diagnosis** | **Affecting growth** | **Growth parameter impaired** |
| --- | --- | --- | --- |
| **Chapter I Infections (detailed)** | | | |
| A00-A09 | Intestinal infections (for age group > 18 months) | temporarily | Weight, BMI (for 4 weeks) |
| A15-A19 | Tuberculosis | permanently | All (for rest of study period) |
| A20-A28 | Certain zoonotic bacterial diseases | temporarily | Weight, BMI (for 4 weeks) |
| A30-A49 | Other bacterial diseases | temporarily | Weight, BMI (for 4 weeks) |
| A50-A52 | Syphillis | permanently | All (for rest of study period) |
| A53-A64 | Other infections with a predominantly sexual mode of transmission | no |  |
| A65-A69 | Other spirochaetal diseases | no |  |
| A70-A74 | Other diseases caused by chlamydiae | no |  |
| A75-A79 | Rickettsioses | temporarily | Weight, BMI (for 4 weeks) |
| A80-A89 | Viral infections of the central nervous system | temporarily | Weight, BMI (for 4 weeks) |
| A92-A99 | Arthropod-borne viral fevers and viral haemorrhagic fevers | temporarily | Weight, BMI (for 4 weeks) |
| B00-B09 | Viral infections characterized by skin and mucous membrane lesions | no |  |
| B15-B17 | Acute viral hepatitis | temporarily | Weight, BMI (for 4 weeks) |
| B18-B19 | Chronic viral hepatitis | permanently | All (for rest of study period) |
| B20-B24 | Human immunodeficiency virus [HIV] disease | permanently | All (for rest of study period) |
| B25-B34 | Other viral diseases | no |  |
| B35-B36 | Mycoses of skin | no |  |
| B37-B46 | Systematic mycoses | temporarily | Weight, BMI (for 4 weeks) |
| B47-B49 | Other mycoses | no |  |
| B50-B64 | Protozoal diseases | temporarily | Weight, BMI (for 4 weeks) |
| B65-B83 | Helminthiases | temporarily | Weight, BMI (for 4 weeks) |
| B85-B99 | Other infectious diseases | no |  |
| **Chapter II Neoplasms** | | | |
| C00-C97 | Malignant neoplasms | permanently | All (for rest of study period) |
| D00-D09 | In situ neoplasms | permanently | All (for rest of study period) |
| D10-D39 | Benign neoplasms in general | no |  |
| D10 | Benign neoplasm of mouth and pharynx | permanently | All (for rest of study period) |
| D11 | Benign neoplasm of major salivary glands | permanently | All (for rest of study period) |
| D12 | Benign neoplasm of colon, rectum, anus and anal canal | permanently | All (for rest of study period) |
| D13 | Benign neoplasm of other and ill-defined parts of digestive system | permanently | All (for rest of study period) |
| D27 | Benign neoplasm of ovary | permanently | All (for rest of study period) |
| D29.2 | Benign neoplasm of male genital organs (Testis = D29.2) | permanently | All (for rest of study period) |
| D30 | Benign neoplasm of urinary organs | permanently | All (for rest of study period) |
| D33 | Benign neoplasm of brain and other parts of central nervous system | permanently | All (for rest of study period) |
| D34 | Benign neoplasm of thyroid gland | permanently | All (for rest of study period) |
| D35 | Benign neoplasm of other and unspecified endocrine glands | permanently | All (for rest of study period) |
| D37-D48 | Neoplasms of uncertain or unknown behaviour | permanently | All (for rest of study period) |
| **Chapter III Diseases of the blood and blood-forming organs** | | | |
| D50-D53 | Nutritional anaemias | no |  |
| D55-D59 | Haemolytic anaemias | permanently | All (for rest of study period) |
| D60-D64 | Aplastic and other anaemias | permanently | All (for rest of study period) |
| D62 | Acute posthaemorrhagic anaemia | no |  |
| D65-D69 | Coagulation defects, purpura and other haemorrhagic conditions | no |  |
| D70-D79 | Other diseases of blood and blood-forming organs | no |  |
| D80-D89 | Certain disorders involving the immune mechanism (Di-George and more) | permanently | All (for rest of study period) |
| **Chapter IV Endocrine, nutritional and metabolic diseases** | | | |
| E00-E07 | Disorders of thyroid gland | permanently | All (for rest of study period) |
| E10-E14 | Diabetes mellitus | permanently | All (for rest of study period) |
| E15-E16 | Other disorders of glucose regulation and pancreatic internal secretion | permanently | All (for rest of study period) |
| E20-E35 | Disorders of other endocrine glands | permanently | All (for rest of study period) |
| E40-E46 | Malnutrition | permanently | All (for rest of study period) |
| E50-E64 | Other nutritional deficiencies | permanently | All (for rest of study period) |
| E65-E68 | Obesity and other hyperalimentation | permanently | All (for rest of study period) |
| E70-E90 | Metabolic disorders | permanently | All (for rest of study period) |
| **Chapter V Mental and behavioural disorders** | | | |
| F00-F99 |  | no |  |
| F10-F19 | Mental and behavioural disorders due to psychoactive substance use | permanently | All (for rest of study period) |
| F32 | Depressive episode | permanently | All (for rest of study period) |
| F33 | Recurrent depressive disorder | permanently | All (for rest of study period) |
| F50 | Eating disorders | permanently | All (for rest of study period) |
| F55 | Abuse of non-dependence-producing substances | permanently | All (for rest of study period) |
| **Chapter VI Diseases of the nervous system** | | | |
| G00-G99 |  | no |  |
| G00-G09 (excl. G08, G09) | Inflammatory diseases of the central nervous system | permanently | All (for rest of study period) |
| G10-G14 | Systemic atrophies primarily affecting the central nervous system | permanently | All (for rest of study period) |
| G60 | Hereditary and idiopathic neuropathy | permanently | All (for rest of study period) |
| G71 | Primary disorders of muscles | permanently | All (for rest of study period) |
| **Chapter VII Diseases of the eye and adnexa** | | | |
| H00-H59 |  | no |  |
| **Chapter VIII Diseases of the ear and mastoid process** | | | |
| H60-H95 |  | no |  |
| **Chapter IX Diseases of the circulatory system** | | | |
| I00-I99 |  | no |  |
| I00-I02 | Acute rheumatic fever | temporarily | Weight, BMI (for 4 weeks) |
| I05-I09 | Chronic rheumatic heart diseases | permanently | All (for rest of study period) |
| I30 | Acute pericarditis | temporarily | Weight, BMI (for 4 weeks) |
| I33 | Acute and subacute endocarditis | temporarily | Weight, BMI (for 4 weeks) |
| I40 | Acute myocarditis | temporarily | Weight, BMI (for 4 weeks) |
| **Chapter X Diseases of the respiratory system** | | | |
| J00-J06 | Acute upper respiratory infections | no |  |
| J09-J18 | Influenza and pneumonia | temporarily | Weight, BMI (for 4 weeks) |
| J20-J22 | Other acute lower respiratory infections | temporarily | Weight, BMI (for 4 weeks) |
| J30 | Vasomotor and allergic rhinitis | no |  |
| J31 | Chronic rhinitis, nasopharyngitis and pharyngitis | no |  |
| J32 | Chronic sinusitis | no |  |
| J35 | Chronic diseases of tonsils and adenoids | no |  |
| J36 | Peritonsillar abscess | no |  |
| J40-J47 | Chronic lower respiratory diseases (Asthma) | no |  |
| J60-J70 | Lung diseases due to external agents | no |  |
| J80-J84 | Other respiratory diseases principally affecting the interstitium | no |  |
| J85-J86 | Suppurative and necrotic conditions of lower respiratory tract | no |  |
| J90-J99 | Other diseases of the pleural or respiratory system | no |  |
| **Chapter XI Diseases of the digestive system** | | | |
| K00-K14 | Diseases of oral cavity, salivary glands and jaws | no |  |
| K20-K31 | Diseases of oesophagus, stomach and duodenum | no |  |
| K35-K37 | Appendicitis | temporarily | Weight, BMI (for 4 weeks) |
| K38 | Other diseases of appendix | no |  |
| K40-K46 | Hernia | no |  |
| K50-K52 | Noninfective enteritis and colitis | temporarily | Weight, BMI (for 4 weeks) |
| K55-K64 | Other diseases of intestines | no |  |
| K65 | Peritonitis | temporarily | Weight, BMI (for 4 weeks) |
| K66-K67 | Other disorders of peritoneum | no |  |
| K70-K77 | Diseases of liver | permanently | All (for rest of study period) |
| K81 | Cholecystitis | temporarily | Weight, BMI (for 4 weeks) |
| K85 | Acute pancreatitis | temporarily | Weight, BMI (for 4 weeks) |
| K80-K87 | Other disorders of gallbladder, biliary tract and pancreas | no |  |
| K90 | Intestinal malabsorption | temporarily | Weight, BMI (for 4 weeks) |
| K91 | Postprocedural disorders of digestive system, not elsewhere classified | temporarily | Weight, BMI (for 4 weeks) |
| K92-K93 | Other diseases of digestive system | no |  |
| **Chapter XII Diseases of the skin and subcutaneous tissue** | | | |
| L00 | Staphylococcal scalded skin syndrome | permanently | Weight, BMI (for 4 weeks) |
| L01-L02 | Impetigo, cutaneous abscess, furuncle and carbuncle | no |  |
| L03-L05 | Cellulitis, Acute lymphadenitis, Pilonidal cyst | no |  |
| L08 | Other local infections of skin and subcutaneous tissue | no |  |
| L10-L14 | Bullous disorders | no |  |
| L20-L30 | Dermatitis and eczema | no |  |
| L40-L45 | Papulosquamous disorders | no |  |
| L50-L54 | Urticaria and erythema | no |  |
| L55-L59 | Radiation-related disorders of the skin and subcutaneous tissue | no |  |
| L60-L75 | Disorders of skin appendages | no |  |
| L80-L99 | Other disorders of the skin and subcutaneous tissue | no |  |
| **Chapter XIII Diseases of the musculoskeletal system and connective tissue** | | | |
| M00-M03 | Infectious arthropathies | temporarily | Weight, BMI (for 4 weeks) |
| M30-M36 | Systemic connective tissue disorders (glucocorticoid treatment) | permanently | All (for rest of study period) |
| M40-M54 | Dorsopathies | no |  |
| M60-M79 | Soft tissue disorders | no |  |
| M80-M85 | Disorders of bone density and structure | no |  |
| M86-M90 | Other osteopathies | permanently | All (for rest of study period) |
| M91-M94 | Chondropathies | permanently | All (for rest of study period) |
| M95-M99 | Other disorders of the musculoskeletal system and connective tissue | no |  |
| **Chapter XIV Diseases of the genitourinary system** | | | |
| N00-N08 | Glomerular diseases | permanently | All (for rest of study period) |
| N10-N16 | Renal tubulo-interstitial diseases | permanently | All (for rest of study period) |
| N17-N19 | Renal failure | permanently | All (for rest of study period) |
| N20-N23 | Urolithiasis | no |  |
| N25-N29 | Other disorders of kidney and ureter | permanently | All (for rest of study period) |
| N30-N39 | Other diseases of urinary system | no |  |
| N40-N51 | Diseases of male genital organs | no |  |
| N60-N64 | Disorders of breast | no |  |
| N70-N71 | Salpingitis and oophoritis, Inflammatory disease of uterus, except cervix | temporarily | Weight, BMI (for 4 weeks) |
| N72-N77 | Inflammatory diseases of female pelvic organs | no |  |
| N80-N98 | Noninflammatory disorders of female genital tract | no |  |
| N99 | Other disorders of the genitourinary system | no |  |
| **Chapter XV Pregnancy, childbirth and the puerperium** | | | |
| O00-O99 | Pregnancy | permanently | Weight, BMI (from pregnancy onwards) |
| **Chapter XVI Certain conditions originating in the perinatal period** | | | |
| P00-P04 | Fetus and newborn affected by maternal factors and by complications of pregnancy, labour and delivery | temporarily | all (for first 18 months) |
| P05 | Slow fetal growth and fetal malnutrition  🡪 Small for gestational age? | temporarily | all (for first 18 months) |
| P07 | Disorders related to short gestation and low birth weight, not elsewhere classified  🡪 Low birth weight? | temporarily | all (for first 18 months) |
| P08 | Disorders related to long gestation and high birth weight | no |  |
| P10-P15 | Birth trauma | no |  |
| P20-P29 | Respiratory and cardiovascular disorders specific to the perinatal period | temporarily | all (for first 18 months) |
| P35-P39 | Infections specific to the perinatal period | temporarily | all (for first 18 months) |
| P51 | Umbilical haemorrhage of newborn | no |  |
| P52 | Intracranial nontraumatic haemorrhage of fetus and newborn | temporarily | all (for first 18 months) |
| P53-P55 | Haemorrhagic disease, haemolytic disease | no |  |
| P56 | Hydrops fetalis due to haemolytic disease | temporarily | all (for first 18 months) |
| P57-P61 | Kernicterus, jaundice, other | no |  |
| P70-P74 | Transitory endocrine and metabolic disorders specific to fetus and newborn | temporarily | all (for first 18 months) |
| P75-P78 | Digestive system disorders of fetus and newborn | temporarily | all (for first 18 months) |
| P80-P83 | Conditions involving the integument and temperature regulation of fetus and newborn | no |  |
| P90-P96 | Other disorders originating in the perinatal period  (P92 Feeding problems?) | no |  |
| **Chapter XVII Congenital malformations, deformations and chromosomal abnormalities** | | | |
| Q00-Q07 | Congenital malformations of the nervous system | permanently | All (for whole study period) |
| Q10-Q18 | Congenital malformations of eye, ear, face and neck | no |  |
| Q20-Q28 | Congenital malformations of the circulatory system | permanently | All (for whole study period) |
| Q30-Q31 | Congenital malformations of nose, larynx | no |  |
| Q32-Q33 | Congenital malformations of trachea, lung | permanently | All (for whole study period) |
| Q34 | Other congenital malformations of respiratory system | no |  |
| Q35-Q37 | Cleft lip and cleft palate | no |  |
| Q38-Q45 | Other congenital malformations of the digestive system | permanently | All (for whole study period) |
| Q50-Q55 | Congenital malformations of genital organs | no |  |
| Q56 | Indeterminate sex and pseudohermaphroditism | permanently | All (for whole study period) |
| Q60-Q64 | Congenital malformations of the urinary system | permanently | All (for whole study period) |
| Q65-Q79 | Congenital malformations and deformations of the musculoskeletal system | permanently | All (for whole study period) |
| Q80-Q85 | Other congenital malformations | no |  |
| Q86-Q89 | Congenital malformation syndromes | permanently | All (for whole study period) |
| Q90-Q99 | Chromosomal abnormalities, not elsewhere classified | permanently | All (for whole study period) |
| **Chapter XVIII Symptoms, signs and abnormal clinical and laboratory findings, not elsewhere classified** | | | |
| R00-R99 |  | no |  |
| **Chapter XIX Injury, poisoning and certain other consequences of external causes** | | | |
| S00-S99 |  | no |  |
| S12 | Fracture of neck | permanently | Height (since injury) |
| S22 | Fracture of rib(s), sternum and thoracic spine | permanently | Height (since injury) |
| S72 | Fracture of femur | permanently | Height (since injury) |
| S78 | Traumatic amputation of hip and thigh | permanently | Height, weight (since injury) |
| S82 | Fracture of lower leg, including ankle | permanently | Height (since injury) |
| S88 | Traumatic amputation of lower leg | permanently | Height, weight (since injury) |
| S92 | Fracture of foot, except ankle | permanently | Height (since injury) |
| S98 | Traumatic amputation of ankle and foot | permanently | Height, weight (since injury) |
| T00-T98 | Trauma (multiple body regions) | no |  |
| **Chapter XX External causes of morbidity and mortality** | | | |
| V01-Y98 |  | no |  |
| Y83.5 | Amputation of limb(s) | permanently | Height, weight (since injury) |
| **Chapter XXI Factors influencing health status and contact with health services** | | | |
| Z00-Z99 |  | no |  |
| Chapter XXII Codes for special purposes | | | |
| U00-U85 |  | no |  |

The categorization of the ICD-10 codes is based on exclusion criteria of previous growth studies and expert opinion [9, 10].
Abbreviations: BMI: Body mass index; ICD-10: International classification of diseases 10th version

**Table S3. Most common nationalities of patients in the SwissPedGrowth project**.

|  | **Patients with available nationality,** n (%)  465,865 (97) |
| --- | --- |
| **Nationality**, n (%) |  |
| Switzerland | 30,820 (71) |
| Germany | 19,205 (4) |
| Portugal | 14,870 (3) |
| Italy | 11,716 (3) |
| France | 10,081 (2) |
| Spain | 5,611 (1) |
| Turkey | 5,362 (1) |
| Eritrea | 5,305 (1) |
| North Macedonia | 3,923 (<1) |
| Serbia | 3,638 (<1) |
| Syrian Arab Republic | 3,571 (<1) |
| Sri Lanka | 2,457 (<1) |
| Ukraine | 2,443 (<1) |
| Afghanistan | 2,188 (<1) |
| Poland | 2,176 (<1) |
| United States of America | 2,018 (<1) |
| Austria | 1,742 (<1) |
| United Kingdom | 1,540 (<1) |
| Brazil | 1,502 (<1) |
| India | 1,450 (<1) |
| Romania | 1,411 (<1) |
| Somalia | 1,368 (<1) |
| Netherlands | 1,312 (<1) |
| Albania | 1,287 (<1) |
| Hungary | 1,239 (<1) |
| Iraq | 1,204 (<1) |
| Greece | 1,200 (<1) |
| China | 1,1175 (<1) |
| Russian Federation | 1,167 (<1) |
| Croatia | 1,117 (<1) |

**Table S4.** **Availability of anthropometric data per hospital visit in electronic health records of SwissPedGrowth hospitals**.

|  | **Total visits,** n (%)  2,171,633 (100) | **Outpatient visits,** n (%)  1,271,392 (59) | **Emergency visits,** n (%)  707,654 (33) | **Inpatient stays,** n (%)  192,587 (9) |
| --- | --- | --- | --- | --- |
| **Height**, n (%) |  |  |  |  |
| No recording | 1,730,440 (80) | 948,270 (75) | 699,190 (99) | 82,980 (43) |
| One recording | 333,627 (15) | 270,470 (21) | 5,986 (<1) | 57,171 (30) |
| Multiple recordings | 107,566 (5) | 52,652 (4) | 2,478 (<1) | 52,436 (27) |
| Median [IQR]^1^ | 1 [1, 1] | 1 [1, 1] | 1 [1, 2] | 1 [1, 2] |
| **Weight**, n (%) |  |  |  |  |
| No recording | 1,233,518 (57) | 905,450 (71) | 300,810 (43) | 27,258 (14) |
| One recording | 691,660 (32) | 305,553 (24) | 330,679 (47) | 55,428 (29) |
| Multiple recordings | 246,455 (11) | 60,389 (5) | 76,165 (11) | 109,901 (57) |
| Median [IQR]^1^ | 1 [1, 2] | 1 [1, 1] | 1 [1, 1] | 2 [1, 4] |
| **Head circumference**, n (%) |  |  |  |  |
| No recording | 2,050,341 (94) | 1,204,287 (95) | 704,439 (100) | 141,615 (74) |
| One recording | 94,241 (4) | 60,652 (5) | 2,418 (<1) | 31,171 (16) |
| Multiple recordings | 27,051 (1) | 6,453 (<1) | 797 (<1) | 19,801 (10) |
| Median [IQR]^1^ | 1 [1, 1] | 1 [1, 1] | 1 [1, 1] | 1 [1, 2] |
| **BMI^2^**, n (%) |  |  |  |  |
| No value | 1,673,279 (77) | 936,365 (74) | 664,101 (94) | 72,813 (38) |
| One value | 342,219 (16) | 279,543 (22) | 29,786 (4) | 32,890 (17) |
| Multiple values | 156,135 (7) | 55,484 (4) | 13,767 (2) | 86,884 (45) |
| Median [IQR]^1^ | 1 [1, 2] | 1 [1, 1] | 1 [1, 2] | 3 [1, 5] |

^1^ Median [IQR]: We report the median and interquartile range of the number of recordings per visit for visits with one or multiple recordings.

^2^ BMI: We calculated body mass index from weight and height measurements.

Abbreviations: BMI: Body mass index; IQR: Inter quartile range

**Table S5. Comparison of patient characteristics and administrative information of hospital visits with and without a height recording in the SwissPedGrowth project**.

|  | **Height available,** n (%)  441,193 (20) | **No height,** n (%)  1,730,440 (80) | **P Value^1^** |
| --- | --- | --- | --- |
| **Sex**, n (%) |  |  | <0.001 |
| Female | 200,843 (21) | 767,652 (79) |  |
| Male | 240,350 (20) | 962,788 (80) |  |
| **Age at hospital visit (years)** |  |  |  |
| Median [IQR] | 6.6 [1.9, 12.2] | 6.3 [2.3, 11.7] | <0.001 |
| Age group, n (%) |  |  | <0.001 |
| Infancy: <1 | 80,737 (27) | 221,958 (73) |  |
| Preschool age: 1–5 | 127,443 (17) | 619,628 (83) |  |
| School age: 6–11 | 117,951 (20) | 479,385 (80) |  |
| Early adolescence: 12–15 | 88,379 (22) | 306,911 (78) |  |
| Late Adolescence: 16–19 | 26,683 (21) | 102,558 (79) |  |
| **Nationality**, n (%) |  |  | <0.001 |
| Swiss | 308,263 (21) | 1,155,872 (79) |  |
| European | 81,582 (19) | 351,301 (81) |  |
| Non-European | 41,509 (19) | 180,733 (81) |  |
| *Missing* | *9,839* | *42,534* |  |
| **Swiss SEP^2^** |  |  |  |
| Mean (SD) | 64.4 (11.0) | 64.9 (11.0) | <0.001 |
| Quintiles, n (%) |  |  | <0.001 |
| 1 | 73,487 (20) | 286,049 (80) |  |
| 2 | 69,942 (20) | 273,548 (80) |  |
| 3 | 80,170 (20) | 311,525 (80) |  |
| 4 | 79,116 (20) | 320,716 (80) |  |
| 5 | 95,431 (19) | 416,316 (81) |  |
| *Missing* | *43,047* | *122,286* |  |
| **General consent^3^**, n (%) |  |  | <0.001 |
| Accepted | 270,627 (21) | 995,810 (79) |  |
| Informed | 51,606 (22) | 180,687 (78) |  |
| Unknown | 118,960 (18) | 553,943 (82) |  |
| **Type of visit,** n (%) |  |  | <0.001 |
| Outpatient visit | 323,122 (25) | 948,270 (75) |  |
| Emergency visit | 8,464 (1) | 699,190 (99) |  |
| Inpatient stay | 109,607 (57) | 82,980 (43) |  |

^1^ P Value: We used Pearson’s chi squared test for categorical, t-test for normally distributed, and Wilcoxon’s rank sum test for non-normally distributed variables.

^2^ Swiss-SEP: Swiss neighbourhood index of socioeconomic position [2].

^3^ General consent: Patients visiting a Swiss hospital are presented with a general consent form, that allows the hospital to re-use patient data for research purposes. A patient’s general consent status can be “accepted” if the patient accepted and signed the general consent form, “rejected” if the patient rejected and returned the general consent form, “informed” if the patient received the general consent form but did not return it yet, or “unknown” if none of the above was recorded.

Abbreviations: IQR: inter quartile range; SD: Standard deviation

**Table S6. Comparison of patient characteristics and administrative information of hospital visits with and without a weight recording in the SwissPedGrowth project**.

|  | **Weight available,** n (%)  938,115 (43) | **No weight,** n (%)  1,233,518 (57) | **P Value^1^** |
| --- | --- | --- | --- |
| **Sex**, n (%) |  |  | <0.001 |
| Female | 423,266 (44) | 545,229 (56) |  |
| Male | 514,849 (43) | 688,289 (57) |  |
| **Age at hospital visit (years)** |  |  |  |
| Median [IQR] | 5.1 [1.6, 10.5] | 7.3 [2.9, 12.6] | <0.001 |
| Age group, n (%) |  |  | <0.001 |
| Infancy: <1 | 168,079 (56) | 134,616 (44) |  |
| Preschool age: 1–5 | 347,749 (47) | 399,322 (53) |  |
| School age: 6–11 | 242,411 (41) | 354,925 (59) |  |
| Early adolescence: 12–15 | 145,571 (37) | 249,719 (63) |  |
| Late Adolescence: 16–19 | 34,305 (27) | 94,936 (73) |  |
| **Nationality**, n (%) |  |  | <0.001 |
| Swiss | 645,225 (44) | 818,910 (56) |  |
| European | 178,469 (41) | 254,414 (59) |  |
| Non-European | 91,042 (41) | 131,200 (59) |  |
| *Missing* | *23,379* | *28,994* |  |
| **Swiss SEP^2^** |  |  |  |
| Mean (SD) | 64.8 (10.9) | 64.8 (11.0) | 0.681 |
| Quintiles, n (%) |  |  | <0.001 |
| 1 | 151,876 (42) | 207,660 (58) |  |
| 2 | 151,554 (44) | 191,936 (56) |  |
| 3 | 177,582 (45) | 214,113 (55) |  |
| 4 | 173,803 (43) | 226,029 (57) |  |
| 5 | 219,231 (43) | 292,516 (57) |  |
| *Missing* | *65,444* | *101,777* |  |
| **General consent^3^**, n (%) |  |  | <0.001 |
| Accepted | 510,758 (40) | 755,679 (60) |  |
| Informed | 129,643 (56) | 102,650 (44) |  |
| Unknown | 297,714 (44) | 375,189 (56) |  |
| **Type of visit,** n (%) |  |  | <0.001 |
| Outpatient visit | 365,942 (29) | 905,450 (71) |  |
| Emergency visit | 406,844 (57) | 300,810 (43) |  |
| Inpatient visit | 165,329 (86) | 27,258 (14) |  |

^1^ P Value: We used Pearson’s chi squared test for categorical, t-test for normally distributed, and Wilcoxon’s rank sum test for non-normally distributed variables.

^2^ Swiss-SEP: Swiss neighbourhood index of socioeconomic position [2].

^3^ General consent: Patients visiting a Swiss hospital are presented with a general consent form, that allows the hospital to re-use patient data for research purposes. A patient’s general consent status can be “accepted” if the patient accepted and signed the general consent form, “rejected” if the patient rejected and returned the general consent form, “informed” if the patient received the general consent form but did not return it yet, or “unknown” if none of the above was recorded.

Abbreviations: IQR: inter quartile range; SD: Standard deviation

**Table S7.** **Availability of anthropometric data per patient in electronic health records of SwissPedGrowth hospitals**.

|  | **SwissPedGrowth patients,** n (%)  477,531 (100) |
| --- | --- |
| **Height**, n (%) |  |
| No recording | 297,676 (62) |
| One recording | 69,164 (14) |
| Multiple recordings | 110,691 (23) |
| Median [IQR]^1^ | 2 [1, 4] |
| **Weight**, n (%) |  |
| No recording | 116,035 (24) |
| One recording | 127,664 (27) |
| Multiple recordings | 233,832 (49) |
| Median [IQR]^1^ | 2 [1, 4] |
| **Head circumference**, n (%) |  |
| No recording | 408,813 (86) |
| One recording | 34,398 (7) |
| Multiple recordings | 34,320 (7) |
| Median [IQR]^1^ | 1 [1, 3] |
| **BMI^2^**, n (%) |  |
| No value | 298,137 (62) |
| One value | 51,510 (11) |
| Multiple values | 127,884 (27) |
| Median [IQR]^1^ | 3 [1, 6] |

^1^ Median [IQR]: We report the median and interquartile range of the number of recordings per visit for visits with one or multiple recordings.

^2^ BMI: We calculated body mass index from weight and height measurements.

Abbreviations: BMI: Body mass index; IQR: Inter quartile range

**Table S8.** **Quality of the height and weight recordings extracted from electronic health records of SwissPedGrowth hospitals using our self-developed algorithm.**

|  | **Height**, n (%)  675,665 (100) | **Weight**, n (%)  1,853,452 (100) |
| --- | --- | --- |
| **No error**, n (%) |  |  |
| No error | 461,606 (68) | 1,189,983 (68) |
| **Corrected error**, n (%) |  |  |
| Swapped recording; include as other parameter | 994 (<1) | 324 (<1) |
| Corrected unit error | 137 (<1) | 626 (<1) |
| Corrected decimal point error | 10 (<1) | 271 (<1) |
| **Duplicate**, n (%) |  |  |
| Carried forward | 149,254 (22) | 317,788 (18) |
| Same day | 44,233 (7) | 193,316 (11) |
| **Biologically implausible outlier**, n (%) |  |  |
| Z-score outlier | 19,272 (3) | 45,070 (3) |
| **Invalid**, n (%) |  |  |
| Zero or negative recording | 48 (<1) | 33 (<1) |
| Negative age at recording | 111 (<1) | 151 (<1) |

See supplementary methods for details about our self-developed algorithm.

**Table S9.** **Output of the *growthcleanr* package used on height and weight recordings extracted from electronic health records of SwissPedGrowth hospitals.**

| **Growthcleanr output** | **Grouped output** | **Height**, n (%)  675,665 (100) | **Weight**, n (%)  1,853,452 (100) |
| --- | --- | --- | --- |
| Include | No error | 480,524 (71) | 1,298,196 (74) |
| Swapped-Measurements | Swapped measurements | 71 (<1) | 72 (<1) |
| Exclude-Adult-Swapped-Measurements |  | - | - |
| Unit-Error-High | Corrected unit error | - | - |
| Unit-Error-Low |  | - | - |
| Unit-Error-Possible |  | - | - |
| Exclude-Adult-Unit-Errors |  | - | - |
| Exclude-Adult-Unit-Errors-RV |  | - | - |
| Exclude-Adult-Hundreds | Corrected digit error | - | - |
| Exclude-Adult-Transpositions |  | - | - |
| Exclude-Adult-Transpositions-RV |  | - | - |
| Exclude-Carried-Forward | Carried forward | 135,847 (20) | 242,138 (14) |
| Exclude-Temporary-Extraneous-Same-Day | Same day | - | - |
| Exclude-Extraneous-Same-Day |  | 46,465 (7) | 197,360 (11) |
| Exclude-Adult-Identical-Same-Day |  | 56 (<1) | 58 (<1) |
| Exclude-Adult-Extraneous-Same-Day |  | 14 (<1) | 18 (<1) |
| Exclude-SD-Cutoff | Single outlier | 261 (<1) | 1,178 (<1) |
| Exclude-Single-Outlier |  | 667 (<1) | 11,471 (<1) |
| Exclude-Adult-BIV |  | - | 1 (<1) |
| Exclude-Adult-Weight-Cap-Identical |  | NA | - |
| Exclude-Adult-Weight-Cap |  | NA | - |
| Exclude-Adult-Distinct-Single |  | - | - |
| Exclude-EWMA-Extreme | Longitudinal outlier | 1,722 (<1) | 1,181 (<1) |
| Exclude-EWMA-Extreme-Pair |  | 468 (<1) | 302 (<1) |
| Exclude-EWMA-8 |  | 3,125 (<1) | 2,030 (<1) |
| Exclude-EWMA-9 |  | 730 (<1) | 1,094 (<1) |
| Exclude-EWMA-10 |  | - | - |
| Exclude-EWMA-11 |  | 353 (<1) | 600 (<1) |
| Exclude-EWMA-12 |  | 199 (<1) | 141 (<1) |
| Exclude-EWMA-13 |  | 4 (<1) | 35 (<1) |
| Exclude-EWMA-14 |  | - | 6 (<1) |
| Exclude-Min-Height-Change |  | 3,498 (1) | NA |
| Exclude-Max-Height-Change |  | 195 (<1) | NA |
| Exclude-Pair-Delta-17 |  | 71 (<1) | 145 (<1) |
| Exclude-Pair-Delta-18 |  | 516 (<1) | 457 (<1) |
| Exclude-Pair-Delta-19 |  | - | - |
| Exclude-Too-Many-Errors |  | 689 (<1) | 184 (<1) |
| Exclude-Too-Many-Errors-Other-Parameter |  | 47 (<1) | 723 (<1) |
| Exclude-Adult-Distinct-Pairs |  | - | - |
| Exclude-Adult-Distinct-3-Or-More |  | 2 (<1) | - |
| Exclude-Adult-EWMA-Extreme |  | - | 1 (<1) |
| Exclude-Adult-EWMA-Extreme-RV |  | - | 1 (<1) |
| Exclude-Adult-Distinct-Ordered-Pairs |  | - | - |
| Exclude-Adult-EWMA-Moderate |  | - | - |
| Exclude-Adult-Possibly-Impacted-By-Weight-Cap |  | - | - |
| Exclude-Adult-Too-Many-Errors |  | - | - |
| Missing | Invalid | 141 (<1) | 170 (<1) |

Abbreviations: BIV: Biologically implausible value; EWMA: Exponentially weighted moving average; RV: Repeated value; SD: Standard deviation.

**Table S10.** **Quality of the height and weight recordings extracted from electronic health records of SwissPedGrowth hospitals combining our self-developed algorithm and the growthcleanr algorithm.**

|  | **Height**, n (%)  675,665 (100) | **Weight**, n (%)  1,853,452 (100) |
| --- | --- | --- |
| **No error**, n (%) |  |  |
| No error | 451,054 (67) | 1,160,316 (66) |
| **Corrected error**, n (%) |  |  |
| Swapped recording; include as other parameter, *self-developed or growthcleanr* | 420 (<1) | 198 (<1) |
| Corrected unit error, *self-developed or growthcleanr* | 112 (<1) | 599 (<1) |
| Corrected decimal point error, *self-developed algorithm* | 8 (<1) | 180 (<1) |
| **Duplicate**, n (%) |  |  |
| Carried forward, *self-developed or growthcleanr* | 150,177 (22) | 311,150 (18) |
| Same day, *self-developed or growthcleanr* | 45,182 (7) | 222,501 (13) |
| **Biologically implausible outlier**, n (%) |  |  |
| Longitudinal outlier, *growthcleanr* | 11,062 (2) | 6,733 (<1) |
| Single outlier, *growthcleanr* | 656 (<1) | 1,717 (<1) |
| Z-score outlier, *self-developed algorithm* | 16,835 (2) | 43,984 (3) |
| **Invalid**, n (%) |  |  |
| Zero or negative recording, *self-developed or growthcleanr* | 48 (<1) | 33 (<1) |
| Negative age at recording, *self-developed or growthcleanr* | 111 (<1) | 151 (<1) |

See supplementary methods for details about algorithms.

**Table S11.** **Quality of the head circumference recordings extracted from electronic health records of SwissPedGrowth hospitals using our self-developed algorithm.**

|  | **Head circumference**, n (%)  192,631 (100) |
| --- | --- |
| **No error**, n (%) |  |
| No error | 143,486 (74) |
| **Corrected error**, n (%) |  |
| Corrected unit error | - |
| Corrected decimal point error | 19 (<1) |
| **Duplicate**, n (%) |  |
| Carried forward | 31,054 (16) |
| Same day | 3,631 (2) |
| **Biologically implausible outlier**, n (%) |  |
| Z-score outlier | 14,274 (7) |
| **Invalid**, n (%) |  |
| Zero or negative recording | 45 (<1) |
| Negative age at recording | 122 (<1) |

See supplementary methods for details about our self-developed algorithm.

**Table S12.** **Quality of the calculated BMI values from height and weight recordings extracted from electronic health records of SwissPedGrowth hospitals combining our self-developed algorithm and the growthcleanr algorithm.**

|  | **BMI**, n (%)  1,169,075 (100) |
| --- | --- |
| **No error**, n (%) |  |
| No error | 657,613 (56) |
| **Corrected error**, n (%) |  |
| Corrected swapped recording, *height or weight* | 302 (<1) |
| Corrected unit error, *height or weight* | 429 (<1) |
| Corrected decimal point error, *height or weight* | 90 (<1) |
| **Duplicate**, n (%) |  |
| Carried forward, *height or weight* | 292,781 (25) |
| Same day, *height or weight* | 148,743 (13) |
| **Biologically implausible outlier**, n (%) |  |
| Longitudinal outlier, *height or weight* | 11,763 (1) |
| Single outlier, *height or weight* | 1,567 (<1) |
| Z-score outlier, *height or weight* | 53,217 (5) |
| Z-score outlier, *BMI* | 2,413 (<1) |
| **Invalid**, n (%) |  |
| Zero or negative recording, *height or weight* | 27 (<1) |
| Negative age at recording, *height or weight* | 130 (<1) |

We used our self-developed algorithm and the growthcleanr algorithm combined to flag height and weight recordings; we additionally excluded BMI values with z-scores <-5 or >8. See supplementary methods for details about the algorithms.
Abbreviations: BMI: Body mass index

**Table S13. Comparison of patients in SwissPedGrowth visiting hospitals during 2017 with the general population of children and adolescents below 20 years living in Switzerland in 2017**.

|  | **Before weighting** | | | **After weighting^1^** | | |
| --- | --- | --- | --- | --- | --- | --- |
|  | **SwissPedGrowth** n (%)  100,752 (6) | **General population** n (%)  1,721,223 (100) | **Standardized difference^2^** | **SwissPedGrowth** n (%_w_)  85,750 (5) | **General population** n (%)  1,721,223 (100) | **Standardized difference^2^** |
| **Age (years) at end of 2017** |  |  |  |  |  |  |
| Age group, n (%) |  |  |  |  |  |  |
| Infancy: <1 | 10,992 (11) | 86,635 (5) | 0.221 | 9,268 (5) | 86,635 (5) | <0.001 |
| Preschool age: 1–5 | 39,680 (39) | 436,750 (25) | 0.301 | 34,341 (25) | 436,750 (25) | <0.001 |
| School age: 6–11 | 28,444 (28) | 508,527 (30) | -0.029 | 24,244 (30) | 508,527 (30) | <0.001 |
| Early adolescence: 12–15 | 16,194 (16) | 329,213 (19) | -0.080 | 13,592 (19) | 329,213 (19) | <0.001 |
| Late Adolescence: 16–19 | 5,442 (5) | 360,098 (21) | -0.481 | 4,305 (21) | 360,098 (21) | <0.001 |
| **Sex**, n (%) |  |  |  |  |  |  |
| Female | 44,803 (44) | 835,803 (49) | -0.082 | 38,072 (49) | 835,803 (49) | <0.001 |
| Male | 55,949 (56) | 885,420 (51) | 0.082 | 47,678 (51) | 885,420 (51) | <0.001 |
| **Nationality**, n (%) |  |  |  |  |  |  |
| Swiss | 67,572 (69) | 1,275,866 (74) | -0.114 | 60,697 (74) | 1,275,866 (74) | <0.001 |
| European | 20,347 (21) | 350,919 (20) | -0.009 | 16,546 (20) | 350,919 (20) | <0.001 |
| Non-European | 9,994 (10) | 93,432 (5) | 0.180 | 8,507 (5) | 93,432 (5) | <0.001 |
| *Missing* | *2,839 (2)* | *1,006 (<1)* | *-* | *-* | *1,006 (<1)* |  |
| **Swiss SEP^1^** |  |  |  |  |  |  |
| Mean (SD) | 65.2 (11.1) | 62.2 (11.0) | 0.273 | 62.7 (11.0) | 62.2 (11.0) | 0.045 |
| Quintiles, n (%) |  |  |  |  |  |  |
| 1 | 14,984 (17) | 396,547 (23) | -0.151 | 14,317 (23) | 396,547 (23) | <0.001 |
| 2 | 14,486 (16) | 342,146 (20) | -0.089 | 14,019 (20) | 342,146 (20) | <0.001 |
| 3 | 17,375 (20) | 334,214 (19) | 0.007 | 16,901 (19) | 334,214 (19) | <0.001 |
| 4 | 17,629 (20) | 331,938 (19) | 0.018 | 17,229 (19) | 331,938 (19) | <0.001 |
| 5 | 23,684 (27) | 316,378 (18) | 0.204 | 23,284 (18) | 316,378 (18) | <0.001 |
| *Missing* | *12,594 (13)* | *-* | *-* | *-* | *-* |  |

^1^ Weighting: For the weighting, we excluded 3% of the SwissPedGrowth participants with missing Nationality and 12% with missing Swiss-SEP. We used the survey package in R with the raking function to estimate weights for SwissPedGrowth participants.
^2^ Standardized difference: We calculated standardized mean differences using Glass’ Δ for normally distributed variables and calculated standardized proportion differences using Cohen’s h for categorical variables [11, 12]. We interpreted the effect size of the standardized differences between the SwissPedGrowth and General population according to Cohen [12]: 0.2 small, 0.5 medium, 0.8 large.
Abbreviations: SD: Standard deviation

**Table S14. Comparison of patients in SwissPedGrowth visiting hospitals during 2023 with the general population of children and adolescents below 20 years living in Switzerland in 2023**.

|  | **Before weighting** | | | **After weighting^1^** | | |
| --- | --- | --- | --- | --- | --- | --- |
|  | **SwissPedGrowth** n (%)  89,500 (5) | **General population** n (%)  1,818,310 (100) | **Standardized difference^2^** | **SwissPedGrowth** n (%)  82,066 (5) | **General population** n (%)  1,818,310 (100) | **Standardized difference^2^** |
| **Age (years) at end of the year** |  |  |  |  |  |  |
| Age group, n (%) |  |  |  |  |  |  |
| Infancy: <1 | 5,562 (6) | 79,437 (4) | 0.083 | 5,108 (4) | 79,437 (4) | <0.001 |
| Preschool age: 1–5 | 32,922 (37) | 447,025 (25) | 0.266 | 30,290 (25) | 447,025 (25) | <0.001 |
| School age: 6–11 | 28,631 (32) | 555,733 (31) | 0.031 | 26,429 (31) | 555,733 (31) | <0.001 |
| Early adolescence: 12–15 | 15,463 (17) | 366,994 (20) | -0.075 | 14,073 (20) | 366,994 (20) | <0.001 |
| Late Adolescence: 16–19 | 6,922 (8) | 369,121 (20) | -0.371 | 6,166 (20) | 369,121 (20) | <0.001 |
| **Sex**, n (%) |  |  |  |  |  |  |
| Female | 40,126 (45) | 880,759 (48) | -0.072 | 36,793 (48) | 880,759 (48) | <0.001 |
| Male | 49,374 (55) | 937,551 (52) | 0.072 | 45,273 (52) | 937,551 (52) | <0.001 |
| **Nationality**, n (%) |  |  |  |  |  |  |
| Swiss | 61,429 (70) | 1,313,646 (72) | -0.050 | 58,442 (72) | 1,313,646 (72) | <0.001 |
| European | 18,799 (21) | 393,631 (22) | -0.006 | 16,630 (22) | 393,631 (22) | <0.001 |
| Non-European | 7,548 (9) | 109,901 (6) | 0.098 | 6,994 (6) | 109,901 (6) | <0.001 |
| *Missing* | *1,724 (2)* | *1,132 (<1)* | *-* | *-* | *1,132 (<1)* |  |
| **Swiss SEP^1^** |  |  |  |  |  |  |
| Mean (SD) | 65.6 (11.0) | 62.8 (10.9) | 0.257 | 63.0 (10.9) | 63.0 (10.0) | <0.001 |
| Quintiles, n (%) |  |  |  |  |  |  |
| 1 | 13,380 (16) | 405,859 (22) | -0.161 | 12,927 (22) | 405,859 (22) | <0.001 |
| 2 | 13,590 (16) | 360,395 (20) | -0.093 | 13,227 (20) | 360,395 (20) | <0.001 |
| 3 | 16,146 (19) | 356,101 (20) | -0.007 | 15,822 (20) | 356,101 (20) | <0.001 |
| 4 | 16,985 (20) | 352,534 (19) | 0.023 | 16,734 (19) | 352,534 (19) | <0.001 |
| 5 | 23,569 (28) | 343,421 (19) | 0.220 | 23,356 (19) | 343,421 (19) | <0.001 |
| *Missing* | *5,830 (7)* | *-* | *-* | *-* | *-* |  |

^1^ Weighting: For the weighting, we excluded 3% of the SwissPedGrowth participants with missing Nationality and 12% with missing Swiss-SEP. We used the survey package in R with the raking function to estimate weights for SwissPedGrowth participants.
^2^ Standardized difference: We calculated standardized mean differences using Glass’ Δ for normally distributed variables and calculated standardized proportion differences using Cohen’s h for categorical variables [11, 12]. We interpreted the effect size of the standardized differences between the SwissPedGrowth and General population according to Cohen [12]: 0.2 small, 0.5 medium, 0.8 large.
Abbreviations: SD: Standard deviation
